# Evaluating Rural/Urban HPV Vaccine Completion Rates in Iowa After the COVID-19 Pandemic

**DOI:** 10.1101/2025.10.07.25337497

**Authors:** Danya Schaefer, Jason Semprini

## Abstract

**Background:** The HPV vaccine provides parents with an opportunity to significantly decrease their children’s future cancer risk. Unfortunately, children face considerable barriers, including stigma and hesitancy, to completing the HPV vaccine schedule. These barriers have contributed to lower HPV vaccination completion rates in rural communities. Although general vaccine hesitancy grew during the pandemic, whether rising vaccine hesitancy further widened rural HPV vaccination gaps remains unknown. Focusing on Iowa, a state with the fastest rising incidence of HPV-associated cancer, we evaluated if county-level COVID-19 vaccination rates corresponded to county-level changes in rural-urban HPV vaccination completion trends.

**Methods:** With data from the Iowa Department of Health and Human Services (2017-2024), we analyzed annual, county-level sex-stratified HPV vaccination completion rates. Rates were reported as a proportion of the 13-15-year-old population. In addition to evaluating overall trends, we grouped counties by rural/urban status and above/below median COVID-19 vaccination rates. We then constructed population-weighted, two-way fixed effect panel regression models testing if HPV vaccination completion rates changed after year 2020; and whether these changes varied by rurality and COVID-19 vaccination rates.

**Results:** Overall, HPV vaccination completion rates increased 5.5%-points (CI = 4.5, 6.5) after 2020 in females and 7.1%-points (CI = 6.0, 8.1) in males. In females, there was no increase after 2020 in HPV vaccine completion in counties with below median COVID-19 vaccine rates (Urban = -0.2%-points, CI = -3.3, 3.3; Rural = 2.9, CI = -0.8, 6.7). In males, the increase after 2020 in HPV vaccine completion rates were consistent across all counties, but lowest in rural counties regardless of COVID-19 vaccine uptake (Above Median = 0.025, CI = 0.010, 0.042; Below Median = 0.027; CI = 0.011, 0.043).

**Conclusions:** In Iowa, the dynamic post-pandemic HPV vaccination completion trends warrant interventions that address multiple factors driving unique contributors to incomplete HPV vaccination adherence.

## Background

Vaccine hesitancy has been a growing concern for public health systems across the globe, generally, but especially so for Human Papillomavirus (HPV), the world’s most prevalent sexually transmitted infection[1–3], prevention systems in rural America[4–6]. Authorized in the U.S. since 2006, the HPV vaccine can prevent certain high-risk strains of HPV to protect children and adolescents against HPV-associated cancers (i.e., cervical cancer, oropharyngeal cancer)[7–9]. General vaccine hesitancy and access barriers, paired with sexual stigma have created a unique combination of factors slowing progress towards HPV vaccination benchmarks[10]. Rural communities in the U.S. have fallen further behind urban progress towards these goals; a trend that will only intensify the rapid rise of HPV-associated cancers in rural America[11].

Children in America face many barriers to completing their HPV vaccine schedule by (the recommended) age thirteen[12]. Even after adjusting for sociodemographic factors, rural vaccination rates lag behind urban rates for general childhood vaccines[13].Many of these barriers are common to the broader healthcare system, just more pronounced across rural communities[14–17]. For example, being insured and visiting the doctor annually at an established source of care are associated with vaccine completion, but rural populations are more likely to be uninsured and less likely to receive preventative healthcare services at an established or high-quality provider. [18– 20]. Yet, even when compared to other childhood vaccines, parents are four times more hesitant to vaccinate their children against HPV[21]. Male children in rural communities have especially low completion rates, with fewer than one in five completing the recommended HPV vaccination schedule[22].

The growing threat of vaccine hesitancy was further fueled during the COVID-19 pandemic[23–25]. While much of the hesitancy was against COVID-19 vaccines, the growing hesitancy was also observed in other childhood vaccines, with significant heterogeneity by geography and politics[26,27]. Recent research also shows the growing hesitancy during the COVID-19 pandemic may be contributing to lower HPV vaccination completion rates[28,29]. While not necessarily causal, there appears to be a strong, inverse correlation between COVID-19 vaccine intent and HPV vaccine hesitancy[30]. Only one study, however, disaggregated pre/post pandemic changes in HPV vaccination (doses) by rurality[31]. Although this claims-based study may not adequately represent the broader population, the results suggest that the HPV vaccinations in rural adolescent males declined more than any other group during the pandemic[31].

Our study builds upon the efforts of Vu et al. by analyzing population-level data on HPV vaccination completion rates, specifically testing for differential changes between rural and urban counties in Iowa[31]. We focus on Iowa, a state with the fastest rising incidence of HPV-associated cancer (Supplemental Exhibit 1). To disentangle rural geography from COVID-19 fueled vaccine hesitancy, we also test if low uptake of the COVID-19 vaccine contributed to any dynamic changes in HPV vaccine completion rates after the pandemic. By quantifying how HPV vaccination completion rates changed during the pandemic, our results can inform targeted interventions by identifying whether geographic disparities are driven by baseline rural access barriers or by the pandemic-fueled growing vaccine hesitancy (or an interaction of the two). Our results can also provide concrete evidence to anecdotal reports informing public health strategies[32].

## Methods

### Data and Measures

We obtained county□level data on HPV vaccination completion rates among adolescents aged 13–15 years in Iowa for 2017 through 2024 from the Iowa Department of Health and Human Services via the Immunization Registry Information System (IRIS)[33]. While this data is publicly available online, the specific data for this report was generated by and received from staff at Iowa’s Department of Health and Human Services (see acknowledgements). IRIS captures individual immunization records from both public and private providers, calculating the proportion of adolescents with a complete HPV vaccine series (defined by receipt of all recommended doses). Rates were calculated using the IRIS-recorded denominator (i.e., the number of active patient records). The IRIS-based denominators ensured that the rate’s numerator was fully contained within its population denominator (IRIS population), enhancing internal consistency. The county-level reports were accessible through the Iowa Public Health Tracking Portal, which allows filtering by county and year, and made available along with this manuscript (Supplemental Exhibit 2).

All HPV vaccination completion rates, reported as a binary variable as a proportion of total 13-15-year-olds, were stratified by sex. Using 2013 Rural Urban Continuum Codes (RUCC), counties were grouped into urban (RUCC = 1-3) or rural (RUCC = 4-9) counties[34]. Counties were also grouped by above/below median 2021 COVID-19 vaccination rates (56.1%)[35]. See county group classifications in Supplemental Exhibit 3.1.

### Statistical Analysis

We estimated a series of panel regression models for female and male HPV vaccination completion rates to assess changes after the COVID-19 pandemic (POST = 1 for years after 2020). The first specification included only the post-pandemic indicator. A second specification interacted POST with an indicator for rural county status to test whether post-2020 changes differed between rural and urban areas. A third specification interacted POST with an indicator for counties below-median COVID-19 vaccination rates to test whether differences were related to pandemic-era vaccine uptake. A final specification included a three-way interaction between POST, rural status, and below-median COVID-19 vaccination rates, allowing for joint assessment of geographic and COVID-19 vaccine coverage effects. For interaction models, we used linear combinations of coefficients to estimate the marginal effect of the post-2020 period for each subgroup. All regressions were implemented in Stata, weighted by population, and included robust standard errors clustered at the county level.

As a sensitivity check, we conducted two alternative analyses. First, we constructed four separate post-period regression models for each mutually exclusive county classification to test if post-period changes varied over time. Second, we re-estimated our primary regression specifications by excluding years 2019-2020.

### Ethics Approval

This study analyzed publicly available, deidentified secondary data and did not involve any interaction or intervention with individuals; therefore, it does not meet the definition of human subjects research under the Common Rule (45 CFR 46.102).

## Results

### Summary Statistics

Between 2017 and 2019, baseline HPV vaccine completion rates were consistently higher among females than males (Table 1). Overall, HPV vaccine completion was 0.442 for females and 0.396 for males. Urban residents demonstrated higher completion than rural residents for both sexes (urban: 0.456 for females and 0.411 for males; rural: 0.418 and 0.371). Within urban areas, those residing in counties with above-median COVID-19 vaccine rates had similar HPV completion compared with those in below-median areas (0.457 vs. 0.434 among females; 0.411 vs. 0.361 among males). In rural settings, HPV completion rates were higher in above-median COVID-19 vaccine rate areas (0.442 for females, 0.400 for males) compared with below-median areas (0.393 and 0.341). Supplemental Exhibit 3.2 reports the baseline (2017-2019) as well as post-period (2021-2024) HPV vaccine completion rates for each county. Figure 1 visualizes the trends in HPV vaccine completion rates for females and males by rural/urban status and above/below median COVID-19 vaccine uptake.

**Table 1.** Baseline (2017-2019) HPV Vaccination Completion Rates. Table 1 reports the average baseline (2017-2019) county-level HPV vaccine completion rate (age 13-15) for females and males by rural status and median 2021 COVID-19 vaccine rates. Urban = RUCC 1-3; Rural = RUCC 4-9.

| **Group** | **Females** | **Males** |
| --- | --- | --- |
| Overall | 0.442 | 0.396 |
| Urban | 0.456 | 0.411 |
| Rural | 0.418 | 0.371 |
| Urban, Above Median COVID-19 Vaccine Rates | 0.457 | 0.411 |
| Urban, Below Median COVID-19 Vaccine Rates | 0.434 | 0.361 |
| Rural, Above Median COVID-19 Vaccine Rates | 0.442 | 0.400 |
| Rural, Below Median COVID-19 Vaccine Rates | 0.393 | 0.341 |

**Figure 1.**
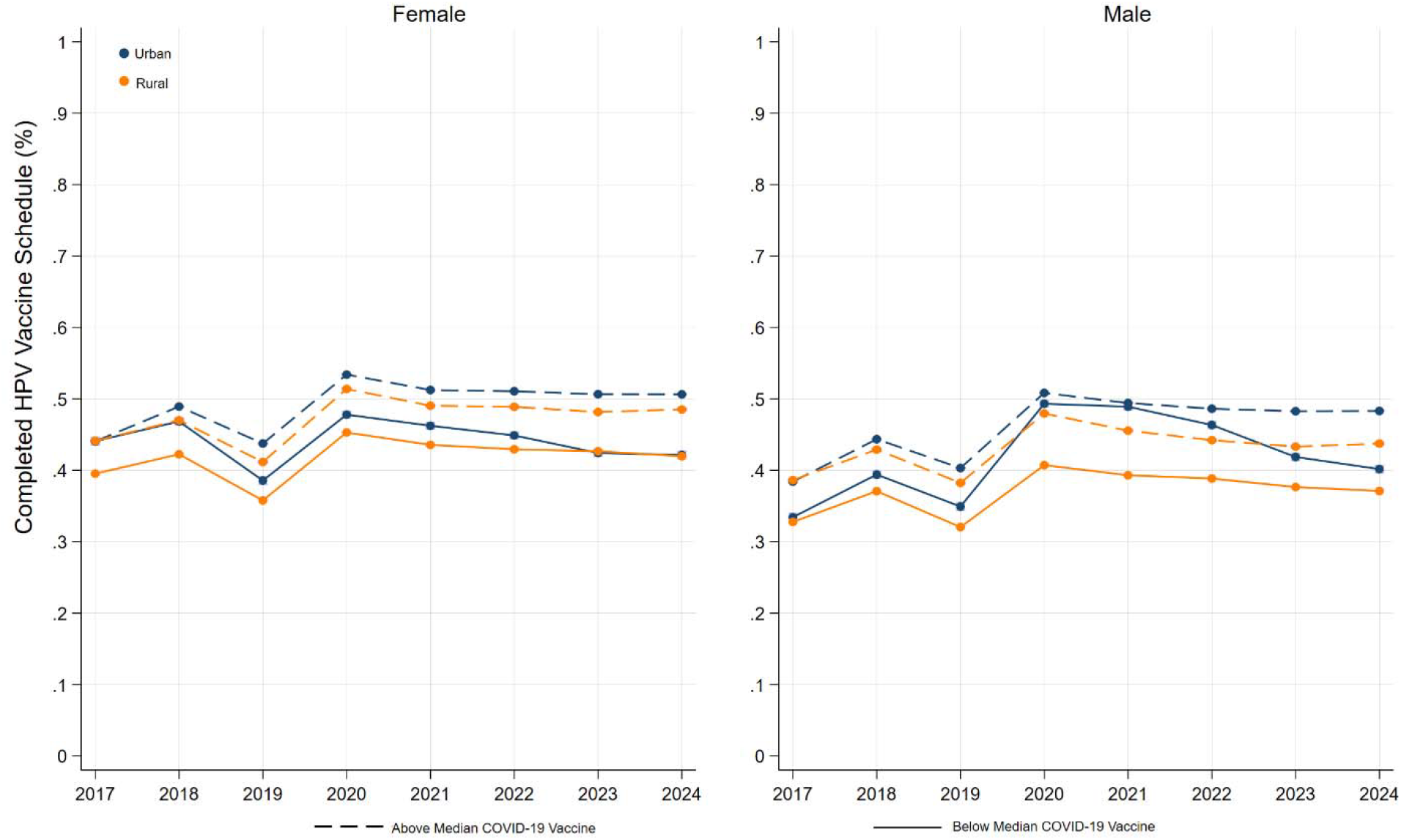
County-Level HPV Vaccine Completion Trends, by sex, rurality, and COVID-19 vaccination rate. Figure 1 reports the average annual county-level HPV vaccine completion rate (age 13-15) for females and males by rural status and median 2021 COVID-19 vaccine rates. Urban = RUCC 1-3; Rural = RUCC 4-9.

### Regression Results

Table 2 reports the results of each regression specification. The overall regression (1) estimated a statistically significant post-period increase in HPV completion rates for females (Est. = 0.031; 95% CI: 0.022, 0.040) and males (Est. = 0.043; 95% CI: 0.034, 0.052). When testing for differences in post-pandemic changes in rural counties (2), we found mixed results by sex. In females, the interaction between POST and rural status was statistically insignificant (Est. = -0.006; 95% CI: –0.020, 0.009). However, in males we the interaction was statistically significant (Est. = –0.024; 95% CI: –0.038, –0.010). When testing for difference in post-pandemic changes by median COVID-19 vaccine uptake, we also saw difference by sex, with only males showing a significant difference from the overall post-period increase (Est. = -0.015; 95% CI = -0.029, -0.000).

**Table 2.** Estimated Association Between COVID-19 Pandemic and HPV Vaccination Completion Rates. Table 2 reports the results of each two-way fixed effect panel regression linear model. Each column was estimated separately. Post = 1 if year > 2020. Rural = 1 if county was a rural county (RUCC 4-7). Below COVID =1 if county reported below median 2021 COVID-19 vaccine rates. 95% confidence intervals reported in brackets. * p<0.05, ** p < 0.01,*** p < 0.001

|  | (1) | (2) | (3) | (4) |
| --- | --- | --- | --- | --- |
|  | **Females** |  |  |  |
| Post | 0.031***<br>[0.022,0.040] | 0.033***<br>[0.020, 0.046] | 0.033***<br>[0.022, 0.043] | 0.034***<br>[0.021,0.046] |
| Post # Rural |  | -0.006<br>[-0.020, 0.009] |  | -0.003<br>[-0.021,0.014] |
| Post # Below<br>COVID |  |  | -0.010<br>[-0.024, 0.003] | -0.035*<br>[-0.045,-0.006] |
| Post # Rural #<br>Below COVID |  |  |  | 0.029<br>[-0.008,0.067] |
|  | **Males** |  |  |  |
| Post | 0.043***<br>[0.034,0.052] | 0.051***<br>[0.041, 0.062] | 0.046***<br>[0.035, 0.056] | 0.051***<br>[0.040,0.062] |
| Post # Rural |  | -0.024**<br>[-0.038, -0.010] |  | -0.026*<br>[-0.045,-0.006] |
| Post # Below<br>COVID |  |  | -0.015*<br>[-0.029, -0.000] | 0.003<br>[-0.010,0.015] |
| Post # Rural #<br>Below COVID |  |  |  | 0.001<br>[-0.021,0.024] |

The final regression specification (4), tested for differential changes across rurality and median COVID-19 vaccine uptake. In females, we estimated a significant interaction between post-period changes and below median COVID-19 vaccine uptake (Est. = -0.035; 95 % CI = -0.045, -0.006). The post-regression analysis estimated that, compared to baseline, there was no change in HPV vaccine completion rates for females in urban counties below the COVID-19 median vaccine rate (Linear Combination Est. = -0.002; 95% CI = -0.033, 0.030) or rural counties below the COVID-19 median vaccine rate (Linear Combination Est. =0.029; CI = -0.008, 0.067). For females, there were no statistically significant differential changes for rural counties. Conversely, in males, we found a statistically significantly different difference for rural counties (Est. = -0.026; 95% CI = -0.045, -0.006). This corresponded to change from baseline for rural counties with above median COVID-19 vaccine rates (Linear Combination Est. = 0.025; 95% CI = 0.010, 0.042) and below median COVID-19 vaccine rates (Linear Combination Est. = 0.027; CI = 0.011, 0.043). There were no statistically significant differences for counties with below COVID-19 vaccine rates for males.

### Alternate Analyses

Supplemental Exhibits 3.3 and 3.4 report the year-by-year post-period coefficients. For females, there were no differences across years for any of the county interaction coefficients. Neither were there any differences across years for males in counties below median COVID-19 vaccine rates. However, we did find evidence that the increase, from baseline years, in HPV vaccine completion rates declined overtime for males in both urban counties above median COVID-19 vaccine rates (Wald test p =0.0036) and rural counties above median COVID-19 vaccine rates (Wald test p = 0.0129). Finally, when excluding 2019 and 2020, our estimates and inference are generally consistent across each specification (Supplemental Exhibit 3.5-7).

## Discussion

Our analysis of population level data in Iowa revealed that HPV vaccine completion rates increased significantly for both females and males from 2017-2024. However, the influence of contextual factors amidst rising HPV vaccination completion rates varied by sex. For females, the overall increase was observed in both rural and urban counties, but remained stagnate after 2020 for females in counties with below-median COVID-19 vaccination rates. For males, the overall increase in HPV vaccination rates did not vary by COVID-19 vaccination rates, but rather by rural-urban status. The observed increase in males was much smaller in rural compared to urban counties.

These results were consistent evidence revealing lower HPV vaccination growth in rural populations[22,36]. Our work builds upon existing studies by showing the heterogeneity by sex, revealing that, at least in Iowa before and after the COVID-19 pandemic, the rural-urban disparity in HPV vaccination completion rate growth is concentrated in males. Future research should investigate whether this heterogeneity generalizes beyond the state of Iowa or if other contextual factors may be mediating the diverging inequities between rural and urban males.

Conversely, when adjusting for COVID-19 vaccination rates, we found no rural-urban disparity in HPV vaccine completion rate growth among females. This result suggests that, unlike the situation with rural males, the factors contributing to stagnate HPV vaccination rates were broader than a single vaccine. These factors could include general vaccine hesitancy within the population, access barriers, or experiencing pandemic related disruptions[37,38]. While future research could illuminate the causes of reduced HPV vaccine completion rates in areas with COVID-19 vaccine rates, our finding reaffirms the evidence that in females barriers to HPV vaccination may mirror barriers to other vaccinations[39].

Interventions to increase HPV vaccination completion will be more effective if tailored to a specific population and designed for contextual factors[40–44]. Our study suggests that, even in a small, relatively homogenous state like Iowa, a blanket approach to driving HPV vaccine completion rates would not necessarily be effective; even if we considered different approaches for rural counties. Instead, leveraging the information of unique contextual factors in females and males could inform more effective strategies. For instance, in females, we need to first understand what barriers prohibited both HPV vaccine completion and COVID-19 vaccine completion. The intervention(s), not specific to any one region, may focus addressing general vaccine hesitancy at the household level or health system barriers related to access or disruptions. In males, interventions can be more targeted towards HPV vaccination and focused on rural communities. This can include combating stigmas surrounding HPV or educating parents to over come the misconception that the HPV vaccination is for females and not males[45,46]. In addition, adolescent well visits may be less frequent in rural communities, and therefore there may be less opportunities to offer the vaccine[47].

The HPV vaccine has been available as a cancer prevention tool in the US for nearly 20 years. Despite progress, HPV vaccination completion remains uneven and context dependent. The persistent rural-urban disparities coincide with rising burden of rural HPV-associated cancers. Policymakers aiming to improve HPV vaccine completion cannot rely on singular solutions or assume rural populations face uniform barriers. Instead, we need nuanced, evidence-informed interventions to account for the intersection of geography, sex, and broader sociocultural attitudes toward vaccination[48]. In Iowa and beyond, public health systems must seek to rebuild trust, close post-pandemic immunization gaps, and reverse diverging trends in HPV vaccination rates. Only with a commitment to understanding and addressing contextual contributors to HPV vaccination disparities can public health systems fully realize the benefit of the HPV vaccine as a cancer prevention tool for the next generational of children growing up in rural America.

### Limitations

Our study is not without its limitations. First, the analysis relied on county-level aggregate data from the Iowa Immunization Registry Information System (IRIS), which, while comprehensive in capturing both public and private provider records, may still be subject to misclassification or incomplete reporting, particularly if there were inconsistencies in provider submissions. Second, the use of IRIS-defined denominators improves internal consistency between numerators and denominators but does not account for adolescents who have moved into or out of a county during the study period, potentially introducing population denominator bias. Third, our ecological design prohibits causal inferences or assessing individual-level barriers influencing these trends. Fourth, the stratification by rural status and by above/below median COVID-19 vaccination rates used broad categorizations that may mask heterogeneity within groups and do not capture other potentially relevant county-level factors such as socioeconomic status, health care access, or political context.

## Conclusions

The HPV vaccine remains a powerful cancer prevention tool. Yet, children, especially those in rural communities, face considerable barriers to completing the HPV vaccine schedule. Given the rising vaccine hesitancy sentiment fueled during the COVID-19 pandemic, we evaluated HPV vaccination completion trends by rural status and COVID-19 vaccine uptake in a state with troubling HPV-associated cancer trends. Although we found increasing completion rates overall, county-level trends varied by sex, rurality, and COVID-19 vaccine uptake. Effective interventions aiming to reverse vaccine hesitancy and advance rural equity must address unique contributors to low HPV vaccination adherence for all children.

## Supporting information

Supplemental Exhibits 1-3

## Data Availability

The data for this study is available as a supplemental file (Supplemental Exhibit 2).

