## Supplemental Exhibits 1-3 for "Evaluating Rural/Urban HPV Vaccine Completion Rates in Iowa After the COVID-19 Pandemic"

**Appendix**

**Supplemental Exhibit 1 – NAACCR Data – HPV-associated Cancer Trends in Rural-Urban states**

**Supplemental Exhibit 2 – Analytic Data**

**Supplemental Exhibit 3.1 – County Classifications**

**Supplemental Exhibit 3.2 – Pre/Post Pandemic HPV Completion Rates, by county**

**Supplemental Exhibit 3.3 –Post-COVID-19 Pandemic Year-by-Year Changes in HPV Vaccine Completion Rates**

**Supplemental Exhibit 3.4 –Post-COVID-19 Pandemic Year-by-Year Changes in HPV Vaccine Completion Rates**

**Supplemental Exhibit 3.5 – Trends in HPV Vaccine Completion Rates – excluding year 2019-2020**

**Supplemental Exhibit 6 – Estimated Association Between COVID-19 Pandemic and HPV Vaccination Completion Rates (Females) – excluding years 2019-2020**

**Supplemental Exhibit 7 – Estimated Association Between COVID-19 Pandemic and HPV Vaccination Completion Rates (Males) - excluding years 2019-2020**

**Supplemental Exhibit 1 – NAACCR Data – HPV-associated Cancer Trends in Rural-Urban states**

| **HPV-associated cancer incidence rate** | **NAACCR Data (1995-2022)** | | | | | |
| --- | --- | --- | --- | --- | --- | --- |
|  | *Metro* | *Metro* | *Metro* | *Non-Metro* | *Non-Metro* | *Non-Metro* |
|  | *APC* | *Lower CI* | *Upper CI* | *APC* | *Lower CI* | *Upper CI* |
| Alabama | ~ | ~ | ~ | ~ | ~ | ~ |
| Alaska | ~ | ~ | ~ | ~ | ~ | ~ |
| Arizona | 0.4* | 0.3 | 0.6 | ~ | ~ | ~ |
| Arkansas | ~ | ~ | ~ | ~ | ~ | ~ |
| California | -0.2* | -0.3 | 0 | 1.4* | 0.9 | 1.8 |
| Colorado | 1.2* | 0.9 | 1.4 | 1.1* | 0.4 | 1.8 |
| Connecticut | 0.7* | 0.3 | 1 | ~ | ~ | ~ |
| Delaware | 0.9* | 0.5 | 1.4 | ~ | ~ | ~ |
| District of Columbia | ~ | ~ | ~ | ~ | ~ | ~ |
| Florida | 0.3* | 0.1 | 0.5 | 1.0* | 0.4 | 1.5 |
| Georgia | ~ | ~ | ~ | ~ | ~ | ~ |
| Hawaii | ~ | ~ | ~ | ~ | ~ | ~ |
| Idaho | 1.3* | 0.8 | 1.8 | 1.2* | 0.5 | 1.8 |
| Illinois | -0.1 | -0.3 | 0.1 | 1.1* | 0.8 | 1.5 |
| Indiana | ~ | ~ | ~ | ~ | ~ | ~ |
| **Iowa** | **1.7*** | **1.2** | **2.1** | **1.8*** | **1.3** | **2.3** |
| Kansas | ~ | ~ | ~ | ~ | ~ | ~ |
| Kentucky | 0.8* | 0.4 | 1.2 | 1.5* | 1.2 | 1.8 |
| Louisiana | 0.4* | 0.2 | 0.7 | 0.7* | 0.2 | 1.1 |
| Maine | 1.0* | 0.4 | 1.6 | 0.6 | 0 | 1.3 |
| Maryland | ~ | ~ | ~ | ~ | ~ | ~ |
| Massachusetts | ~ | ~ | ~ | ~ | ~ | ~ |
| Michigan | 0.4* | 0.2 | 0.7 | 1.1* | 0.7 | 1.5 |
| Minnesota | 0.9* | 0.6 | 1.3 | 1.5* | 1.2 | 1.8 |
| Mississippi | ~ | ~ | ~ | ~ | ~ | ~ |
| Missouri | ~ | ~ | ~ | ~ | ~ | ~ |
| Montana | ~ | ~ | ~ | ~ | ~ | ~ |
| Nebraska | 0.8* | 0.3 | 1.3 | 1.4* | 0.8 | 2 |
| Nevada | ~ | ~ | ~ | ~ | ~ | ~ |
| New Hampshire | ~ | ~ | ~ | ~ | ~ | ~ |
| New Jersey | -0.1 | -0.3 | 0.1 | ~ | ~ | ~ |
| New Mexico | ~ | ~ | ~ | ~ | ~ | ~ |
| New York | 0 | -0.2 | 0.2 | 1.1* | 0.7 | 1.4 |
| North Carolina | 0.9* | 0.7 | 1.1 | 1.1* | 0.8 | 1.4 |
| North Dakota | ~ | ~ | ~ | ~ | ~ | ~ |
| Ohio | ~ | ~ | ~ | ~ | ~ | ~ |
| Oklahoma | ~ | ~ | ~ | ~ | ~ | ~ |
| Oregon | ~ | ~ | ~ | ~ | ~ | ~ |
| Pennsylvania | 0.7* | 0.5 | 0.9 | 1.1* | 0.7 | 1.5 |
| Puerto Rico | ~ | ~ | ~ | ~ | ~ | ~ |
| Rhode Island | 0.4 | -0.1 | 0.8 | ~ | ~ | ~ |
| South Carolina | ~ | ~ | ~ | ~ | ~ | ~ |
| South Dakota | ~ | ~ | ~ | ~ | ~ | ~ |
| Tennessee | ~ | ~ | ~ | ~ | ~ | ~ |
| Texas | 0.1 | -0.1 | 0.3 | 1.0* | 0.4 | 1.5 |
| Utah | 0.9* | 0.6 | 1.3 | ~ | ~ | ~ |
| Vermont | ~ | ~ | ~ | ~ | ~ | ~ |
| Virginia | ~ | ~ | ~ | ~ | ~ | ~ |
| Washington | 0.8* | 0.6 | 1 | 0.9* | 0.3 | 1.5 |
| West Virginia | 0.9* | 0.4 | 1.4 | 0.6* | 0.2 | 1.1 |
| Wisconsin | 0.8* | 0.5 | 1.2 | 1.1* | 0.6 | 1.6 |
| Wyoming | ~ | ~ | ~ | ~ | ~ | ~ |
| Rates are per 100,000 and age-adjusted to the 2000 US Std Population (19 age groups - Census P25-1130) standard; Confidence intervals are 95% for rates (Tiwari mod) and trends.  Percent changes were calculated using 1 year for each end point; APCs were calculated using weighted least squares method.  ~ Statistic could not be calculated. | | | | | | |

**Supplemental Exhibit 2 – Analytic Data**

| year | county | femalecomplete | malecomplete | fips | administered_dose1_pop_pct | rural | covidvax_belowmed | post | avg_pop |
| --- | --- | --- | --- | --- | --- | --- | --- | --- | --- |
| 2017 | Adair | 0.66129 | 0.637931 | 19001 | 51.3 | 1 | 1 | 0 | 165 |
| 2018 | Adair | 0.726563 | 0.651786 | 19001 | 51.3 | 1 | 1 | 0 | 165 |
| 2019 | Adair | 0.659091 | 0.542857 | 19001 | 51.3 | 1 | 1 | 0 | 165 |
| 2020 | Adair | 0.675497 | 0.62963 | 19001 | 51.3 | 1 | 1 | 1 | 165 |
| 2021 | Adair | 0.620253 | 0.633333 | 19001 | 51.3 | 1 | 1 | 1 | 165 |
| 2022 | Adair | 0.568493 | 0.568493 | 19001 | 51.3 | 1 | 1 | 1 | 165 |
| 2023 | Adair | 0.584615 | 0.551282 | 19001 | 51.3 | 1 | 1 | 1 | 165 |
| 2024 | Adair | 0.566667 | 0.534591 | 19001 | 51.3 | 1 | 1 | 1 | 165 |
| 2017 | Adams | 0.439394 | 0.358209 | 19003 | 57.9 | 1 | 0 | 0 | 77 |
| 2018 | Adams | 0.519481 | 0.42029 | 19003 | 57.9 | 1 | 0 | 0 | 77 |
| 2019 | Adams | 0.382979 | 0.352113 | 19003 | 57.9 | 1 | 0 | 0 | 77 |
| 2020 | Adams | 0.470588 | 0.488095 | 19003 | 57.9 | 1 | 0 | 1 | 77 |
| 2021 | Adams | 0.484536 | 0.530864 | 19003 | 57.9 | 1 | 0 | 1 | 77 |
| 2022 | Adams | 0.51087 | 0.44186 | 19003 | 57.9 | 1 | 0 | 1 | 77 |
| 2023 | Adams | 0.655172 | 0.545455 | 19003 | 57.9 | 1 | 0 | 1 | 77 |
| 2024 | Adams | 0.741935 | 0.643836 | 19003 | 57.9 | 1 | 0 | 1 | 77 |
| 2017 | Allamakee | 0.386819 | 0.286957 | 19005 | 56.7 | 1 | 0 | 0 | 274 |
| 2018 | Allamakee | 0.397695 | 0.341954 | 19005 | 56.7 | 1 | 0 | 0 | 274 |
| 2019 | Allamakee | 0.349162 | 0.297368 | 19005 | 56.7 | 1 | 0 | 0 | 274 |
| 2020 | Allamakee | 0.401662 | 0.400468 | 19005 | 56.7 | 1 | 0 | 1 | 274 |
| 2021 | Allamakee | 0.413105 | 0.409091 | 19005 | 56.7 | 1 | 0 | 1 | 274 |
| 2022 | Allamakee | 0.414169 | 0.354592 | 19005 | 56.7 | 1 | 0 | 1 | 274 |
| 2023 | Allamakee | 0.430894 | 0.333333 | 19005 | 56.7 | 1 | 0 | 1 | 274 |
| 2024 | Allamakee | 0.452663 | 0.323625 | 19005 | 56.7 | 1 | 0 | 1 | 274 |
| 2017 | Appanoose | 0.294118 | 0.138996 | 19007 | 49.5 | 1 | 1 | 0 | 148 |
| 2018 | Appanoose | 0.325301 | 0.239544 | 19007 | 49.5 | 1 | 1 | 0 | 148 |
| 2019 | Appanoose | 0.247863 | 0.19697 | 19007 | 49.5 | 1 | 1 | 0 | 148 |
| 2020 | Appanoose | 0.30888 | 0.346939 | 19007 | 49.5 | 1 | 1 | 1 | 148 |
| 2021 | Appanoose | 0.332061 | 0.289474 | 19007 | 49.5 | 1 | 1 | 1 | 148 |
| 2022 | Appanoose | 0.356877 | 0.283217 | 19007 | 49.5 | 1 | 1 | 1 | 148 |
| 2023 | Appanoose | 0.338776 | 0.239437 | 19007 | 49.5 | 1 | 1 | 1 | 148 |
| 2024 | Appanoose | 0.289916 | 0.231884 | 19007 | 49.5 | 1 | 1 | 1 | 148 |
| 2017 | Audubon | 0.528736 | 0.446809 | 19009 | 59.2 | 1 | 0 | 0 | 111 |
| 2018 | Audubon | 0.561224 | 0.546296 | 19009 | 59.2 | 1 | 0 | 0 | 111 |
| 2019 | Audubon | 0.398148 | 0.463636 | 19009 | 59.2 | 1 | 0 | 0 | 111 |
| 2020 | Audubon | 0.529412 | 0.513274 | 19009 | 59.2 | 1 | 0 | 1 | 111 |
| 2021 | Audubon | 0.552632 | 0.473214 | 19009 | 59.2 | 1 | 0 | 1 | 111 |
| 2022 | Audubon | 0.568 | 0.495413 | 19009 | 59.2 | 1 | 0 | 1 | 111 |
| 2023 | Audubon | 0.566038 | 0.456 | 19009 | 59.2 | 1 | 0 | 1 | 111 |
| 2024 | Audubon | 0.607843 | 0.412698 | 19009 | 59.2 | 1 | 0 | 1 | 111 |
| 2017 | Benton | 0.563055 | 0.443595 | 19011 | 59.8 | 0 | 0 | 0 | 633 |
| 2018 | Benton | 0.611418 | 0.527728 | 19011 | 59.8 | 0 | 0 | 0 | 633 |
| 2019 | Benton | 0.527725 | 0.474517 | 19011 | 59.8 | 0 | 0 | 0 | 633 |
| 2020 | Benton | 0.636861 | 0.622977 | 19011 | 59.8 | 0 | 0 | 1 | 633 |
| 2021 | Benton | 0.595238 | 0.584874 | 19011 | 59.8 | 0 | 0 | 1 | 633 |
| 2022 | Benton | 0.598148 | 0.582301 | 19011 | 59.8 | 0 | 0 | 1 | 633 |
| 2023 | Benton | 0.562832 | 0.575175 | 19011 | 59.8 | 0 | 0 | 1 | 633 |
| 2024 | Benton | 0.577617 | 0.546713 | 19011 | 59.8 | 0 | 0 | 1 | 633 |
| 2017 | Black Hawk | 0.525595 | 0.436849 | 19013 | 62.5 | 0 | 0 | 0 | 3159 |
| 2018 | Black Hawk | 0.559603 | 0.48613 | 19013 | 62.5 | 0 | 0 | 0 | 3159 |
| 2019 | Black Hawk | 0.49723 | 0.433014 | 19013 | 62.5 | 0 | 0 | 0 | 3159 |
| 2020 | Black Hawk | 0.589933 | 0.551974 | 19013 | 62.5 | 0 | 0 | 1 | 3159 |
| 2021 | Black Hawk | 0.566901 | 0.54884 | 19013 | 62.5 | 0 | 0 | 1 | 3159 |
| 2022 | Black Hawk | 0.542502 | 0.543894 | 19013 | 62.5 | 0 | 0 | 1 | 3159 |
| 2023 | Black Hawk | 0.526518 | 0.534138 | 19013 | 62.5 | 0 | 0 | 1 | 3159 |
| 2024 | Black Hawk | 0.526506 | 0.53466 | 19013 | 62.5 | 0 | 0 | 1 | 3159 |
| 2017 | Boone | 0.439834 | 0.436782 | 19015 | 65.7 | 1 | 0 | 0 | 585 |
| 2018 | Boone | 0.501002 | 0.500982 | 19015 | 65.7 | 1 | 0 | 0 | 585 |
| 2019 | Boone | 0.462 | 0.442379 | 19015 | 65.7 | 1 | 0 | 0 | 585 |
| 2020 | Boone | 0.585069 | 0.606481 | 19015 | 65.7 | 1 | 0 | 1 | 585 |
| 2021 | Boone | 0.574871 | 0.60972 | 19015 | 65.7 | 1 | 0 | 1 | 585 |
| 2022 | Boone | 0.532203 | 0.537721 | 19015 | 65.7 | 1 | 0 | 1 | 585 |
| 2023 | Boone | 0.520761 | 0.498246 | 19015 | 65.7 | 1 | 0 | 1 | 585 |
| 2024 | Boone | 0.546875 | 0.446691 | 19015 | 65.7 | 1 | 0 | 1 | 585 |
| 2017 | Bremer | 0.564 | 0.534483 | 19017 | 61.8 | 0 | 0 | 0 | 649 |
| 2018 | Bremer | 0.609615 | 0.558442 | 19017 | 61.8 | 0 | 0 | 0 | 649 |
| 2019 | Bremer | 0.530651 | 0.493482 | 19017 | 61.8 | 0 | 0 | 0 | 649 |
| 2020 | Bremer | 0.652557 | 0.633058 | 19017 | 61.8 | 0 | 0 | 1 | 649 |
| 2021 | Bremer | 0.614437 | 0.607492 | 19017 | 61.8 | 0 | 0 | 1 | 649 |
| 2022 | Bremer | 0.611913 | 0.582803 | 19017 | 61.8 | 0 | 0 | 1 | 649 |
| 2023 | Bremer | 0.57197 | 0.591362 | 19017 | 61.8 | 0 | 0 | 1 | 649 |
| 2024 | Bremer | 0.597633 | 0.564189 | 19017 | 61.8 | 0 | 0 | 1 | 649 |
| 2017 | Buchanan | 0.53 | 0.423372 | 19019 | 55.3 | 1 | 1 | 0 | 510 |
| 2018 | Buchanan | 0.547619 | 0.482283 | 19019 | 55.3 | 1 | 1 | 0 | 510 |
| 2019 | Buchanan | 0.452242 | 0.41527 | 19019 | 55.3 | 1 | 1 | 0 | 510 |
| 2020 | Buchanan | 0.530303 | 0.548571 | 19019 | 55.3 | 1 | 1 | 1 | 510 |
| 2021 | Buchanan | 0.547809 | 0.550285 | 19019 | 55.3 | 1 | 1 | 1 | 510 |
| 2022 | Buchanan | 0.54321 | 0.533981 | 19019 | 55.3 | 1 | 1 | 1 | 510 |
| 2023 | Buchanan | 0.504255 | 0.483235 | 19019 | 55.3 | 1 | 1 | 1 | 510 |
| 2024 | Buchanan | 0.504237 | 0.461856 | 19019 | 55.3 | 1 | 1 | 1 | 510 |
| 2017 | Buena Vista | 0.235484 | 0.182874 | 19021 | 69.5 | 1 | 0 | 0 | 352 |
| 2018 | Buena Vista | 0.265306 | 0.236607 | 19021 | 69.5 | 1 | 0 | 0 | 352 |
| 2019 | Buena Vista | 0.21978 | 0.218126 | 19021 | 69.5 | 1 | 0 | 0 | 352 |
| 2020 | Buena Vista | 0.289552 | 0.268657 | 19021 | 69.5 | 1 | 0 | 1 | 352 |
| 2021 | Buena Vista | 0.30229 | 0.222382 | 19021 | 69.5 | 1 | 0 | 1 | 352 |
| 2022 | Buena Vista | 0.316109 | 0.234807 | 19021 | 69.5 | 1 | 0 | 1 | 352 |
| 2023 | Buena Vista | 0.309341 | 0.253078 | 19021 | 69.5 | 1 | 0 | 1 | 352 |
| 2024 | Buena Vista | 0.330409 | 0.289835 | 19021 | 69.5 | 1 | 0 | 1 | 352 |
| 2017 | Butler | 0.566102 | 0.436709 | 19023 | 59 | 1 | 0 | 0 | 361 |
| 2018 | Butler | 0.617162 | 0.496875 | 19023 | 59 | 1 | 0 | 0 | 361 |
| 2019 | Butler | 0.554817 | 0.455882 | 19023 | 59 | 1 | 0 | 0 | 361 |
| 2020 | Butler | 0.647929 | 0.592262 | 19023 | 59 | 1 | 0 | 1 | 361 |
| 2021 | Butler | 0.614198 | 0.572626 | 19023 | 59 | 1 | 0 | 1 | 361 |
| 2022 | Butler | 0.61875 | 0.554286 | 19023 | 59 | 1 | 0 | 1 | 361 |
| 2023 | Butler | 0.618892 | 0.543131 | 19023 | 59 | 1 | 0 | 1 | 361 |
| 2024 | Butler | 0.647458 | 0.508417 | 19023 | 59 | 1 | 0 | 1 | 361 |
| 2017 | Calhoun | 0.345178 | 0.278947 | 19025 | 60.8 | 1 | 0 | 0 | 173 |
| 2018 | Calhoun | 0.446809 | 0.356021 | 19025 | 60.8 | 1 | 0 | 0 | 173 |
| 2019 | Calhoun | 0.365 | 0.308057 | 19025 | 60.8 | 1 | 0 | 0 | 173 |
| 2020 | Calhoun | 0.441624 | 0.484536 | 19025 | 60.8 | 1 | 0 | 1 | 173 |
| 2021 | Calhoun | 0.458937 | 0.462617 | 19025 | 60.8 | 1 | 0 | 1 | 173 |
| 2022 | Calhoun | 0.518692 | 0.410959 | 19025 | 60.8 | 1 | 0 | 1 | 173 |
| 2023 | Calhoun | 0.494565 | 0.443478 | 19025 | 60.8 | 1 | 0 | 1 | 173 |
| 2024 | Calhoun | 0.517766 | 0.439655 | 19025 | 60.8 | 1 | 0 | 1 | 173 |
| 2017 | Carroll | 0.341176 | 0.241901 | 19027 | 63.1 | 1 | 0 | 0 | 329 |
| 2018 | Carroll | 0.420048 | 0.306488 | 19027 | 63.1 | 1 | 0 | 0 | 329 |
| 2019 | Carroll | 0.301552 | 0.247401 | 19027 | 63.1 | 1 | 0 | 0 | 329 |
| 2020 | Carroll | 0.443478 | 0.351456 | 19027 | 63.1 | 1 | 0 | 1 | 329 |
| 2021 | Carroll | 0.405579 | 0.339181 | 19027 | 63.1 | 1 | 0 | 1 | 329 |
| 2022 | Carroll | 0.389978 | 0.318182 | 19027 | 63.1 | 1 | 0 | 1 | 329 |
| 2023 | Carroll | 0.392116 | 0.329854 | 19027 | 63.1 | 1 | 0 | 1 | 329 |
| 2024 | Carroll | 0.411881 | 0.325301 | 19027 | 63.1 | 1 | 0 | 1 | 329 |
| 2017 | Cass | 0.518152 | 0.479876 | 19029 | 60.3 | 1 | 0 | 0 | 327 |
| 2018 | Cass | 0.567669 | 0.543408 | 19029 | 60.3 | 1 | 0 | 0 | 327 |
| 2019 | Cass | 0.486207 | 0.553055 | 19029 | 60.3 | 1 | 0 | 0 | 327 |
| 2020 | Cass | 0.706081 | 0.702703 | 19029 | 60.3 | 1 | 0 | 1 | 327 |
| 2021 | Cass | 0.651316 | 0.619048 | 19029 | 60.3 | 1 | 0 | 1 | 327 |
| 2022 | Cass | 0.578767 | 0.534247 | 19029 | 60.3 | 1 | 0 | 1 | 327 |
| 2023 | Cass | 0.50173 | 0.468439 | 19029 | 60.3 | 1 | 0 | 1 | 327 |
| 2024 | Cass | 0.458333 | 0.442953 | 19029 | 60.3 | 1 | 0 | 1 | 327 |
| 2017 | Cedar | 0.39759 | 0.411458 | 19031 | 61.8 | 1 | 0 | 0 | 389 |
| 2018 | Cedar | 0.507375 | 0.477654 | 19031 | 61.8 | 1 | 0 | 0 | 389 |
| 2019 | Cedar | 0.455072 | 0.408719 | 19031 | 61.8 | 1 | 0 | 0 | 389 |
| 2020 | Cedar | 0.572115 | 0.539024 | 19031 | 61.8 | 1 | 0 | 1 | 389 |
| 2021 | Cedar | 0.559611 | 0.498829 | 19031 | 61.8 | 1 | 0 | 1 | 389 |
| 2022 | Cedar | 0.550617 | 0.485981 | 19031 | 61.8 | 1 | 0 | 1 | 389 |
| 2023 | Cedar | 0.551813 | 0.5 | 19031 | 61.8 | 1 | 0 | 1 | 389 |
| 2024 | Cedar | 0.58871 | 0.498652 | 19031 | 61.8 | 1 | 0 | 1 | 389 |
| 2017 | Cerro Gordo | 0.447073 | 0.433411 | 19033 | 65.9 | 1 | 0 | 0 | 810 |
| 2018 | Cerro Gordo | 0.461063 | 0.451991 | 19033 | 65.9 | 1 | 0 | 0 | 810 |
| 2019 | Cerro Gordo | 0.405762 | 0.381375 | 19033 | 65.9 | 1 | 0 | 0 | 810 |
| 2020 | Cerro Gordo | 0.498922 | 0.482582 | 19033 | 65.9 | 1 | 0 | 1 | 810 |
| 2021 | Cerro Gordo | 0.455556 | 0.445545 | 19033 | 65.9 | 1 | 0 | 1 | 810 |
| 2022 | Cerro Gordo | 0.474333 | 0.424395 | 19033 | 65.9 | 1 | 0 | 1 | 810 |
| 2023 | Cerro Gordo | 0.455641 | 0.409234 | 19033 | 65.9 | 1 | 0 | 1 | 810 |
| 2024 | Cerro Gordo | 0.45403 | 0.401909 | 19033 | 65.9 | 1 | 0 | 1 | 810 |
| 2017 | Cherokee | 0.387255 | 0.344186 | 19035 | 54.9 | 1 | 1 | 0 | 193 |
| 2018 | Cherokee | 0.436681 | 0.399103 | 19035 | 54.9 | 1 | 1 | 0 | 193 |
| 2019 | Cherokee | 0.354839 | 0.305556 | 19035 | 54.9 | 1 | 1 | 0 | 193 |
| 2020 | Cherokee | 0.462451 | 0.417391 | 19035 | 54.9 | 1 | 1 | 1 | 193 |
| 2021 | Cherokee | 0.444444 | 0.354978 | 19035 | 54.9 | 1 | 1 | 1 | 193 |
| 2022 | Cherokee | 0.439689 | 0.353909 | 19035 | 54.9 | 1 | 1 | 1 | 193 |
| 2023 | Cherokee | 0.494382 | 0.378378 | 19035 | 54.9 | 1 | 1 | 1 | 193 |
| 2024 | Cherokee | 0.5 | 0.450893 | 19035 | 54.9 | 1 | 1 | 1 | 193 |
| 2017 | Chickasaw | 0.461207 | 0.47619 | 19037 | 56.3 | 1 | 0 | 0 | 233 |
| 2018 | Chickasaw | 0.484375 | 0.396364 | 19037 | 56.3 | 1 | 0 | 0 | 233 |
| 2019 | Chickasaw | 0.386282 | 0.34965 | 19037 | 56.3 | 1 | 0 | 0 | 233 |
| 2020 | Chickasaw | 0.536232 | 0.456747 | 19037 | 56.3 | 1 | 0 | 1 | 233 |
| 2021 | Chickasaw | 0.547893 | 0.402214 | 19037 | 56.3 | 1 | 0 | 1 | 233 |
| 2022 | Chickasaw | 0.493976 | 0.384 | 19037 | 56.3 | 1 | 0 | 1 | 233 |
| 2023 | Chickasaw | 0.512097 | 0.383721 | 19037 | 56.3 | 1 | 0 | 1 | 233 |
| 2024 | Chickasaw | 0.529183 | 0.421053 | 19037 | 56.3 | 1 | 0 | 1 | 233 |
| 2017 | Clarke | 0.443925 | 0.346614 | 19039 | 56.5 | 1 | 0 | 0 | 205 |
| 2018 | Clarke | 0.502283 | 0.388889 | 19039 | 56.5 | 1 | 0 | 0 | 205 |
| 2019 | Clarke | 0.40625 | 0.32377 | 19039 | 56.5 | 1 | 0 | 0 | 205 |
| 2020 | Clarke | 0.502008 | 0.405204 | 19039 | 56.5 | 1 | 0 | 1 | 205 |
| 2021 | Clarke | 0.418972 | 0.378676 | 19039 | 56.5 | 1 | 0 | 1 | 205 |
| 2022 | Clarke | 0.48062 | 0.394737 | 19039 | 56.5 | 1 | 0 | 1 | 205 |
| 2023 | Clarke | 0.45749 | 0.353846 | 19039 | 56.5 | 1 | 0 | 1 | 205 |
| 2024 | Clarke | 0.478992 | 0.347328 | 19039 | 56.5 | 1 | 0 | 1 | 205 |
| 2017 | Clay | 0.394886 | 0.383333 | 19041 | 54.3 | 1 | 1 | 0 | 251 |
| 2018 | Clay | 0.398892 | 0.363388 | 19041 | 54.3 | 1 | 1 | 0 | 251 |
| 2019 | Clay | 0.309973 | 0.269841 | 19041 | 54.3 | 1 | 1 | 0 | 251 |
| 2020 | Clay | 0.354054 | 0.295673 | 19041 | 54.3 | 1 | 1 | 1 | 251 |
| 2021 | Clay | 0.350877 | 0.276888 | 19041 | 54.3 | 1 | 1 | 1 | 251 |
| 2022 | Clay | 0.343407 | 0.285714 | 19041 | 54.3 | 1 | 1 | 1 | 251 |
| 2023 | Clay | 0.433022 | 0.318452 | 19041 | 54.3 | 1 | 1 | 1 | 251 |
| 2024 | Clay | 0.432258 | 0.325581 | 19041 | 54.3 | 1 | 1 | 1 | 251 |
| 2017 | Clayton | 0.329154 | 0.27566 | 19043 | 50.3 | 1 | 1 | 0 | 253 |
| 2018 | Clayton | 0.411392 | 0.297994 | 19043 | 50.3 | 1 | 1 | 0 | 253 |
| 2019 | Clayton | 0.387195 | 0.254491 | 19043 | 50.3 | 1 | 1 | 0 | 253 |
| 2020 | Clayton | 0.457726 | 0.359551 | 19043 | 50.3 | 1 | 1 | 1 | 253 |
| 2021 | Clayton | 0.391931 | 0.360215 | 19043 | 50.3 | 1 | 1 | 1 | 253 |
| 2022 | Clayton | 0.366071 | 0.372603 | 19043 | 50.3 | 1 | 1 | 1 | 253 |
| 2023 | Clayton | 0.435821 | 0.379404 | 19043 | 50.3 | 1 | 1 | 1 | 253 |
| 2024 | Clayton | 0.474164 | 0.387879 | 19043 | 50.3 | 1 | 1 | 1 | 253 |
| 2017 | Clinton | 0.347689 | 0.270352 | 19045 | 57.6 | 1 | 0 | 0 | 648 |
| 2018 | Clinton | 0.360294 | 0.297165 | 19045 | 57.6 | 1 | 0 | 0 | 648 |
| 2019 | Clinton | 0.273726 | 0.244403 | 19045 | 57.6 | 1 | 0 | 0 | 648 |
| 2020 | Clinton | 0.33758 | 0.284734 | 19045 | 57.6 | 1 | 0 | 1 | 648 |
| 2021 | Clinton | 0.320755 | 0.29732 | 19045 | 57.6 | 1 | 0 | 1 | 648 |
| 2022 | Clinton | 0.300357 | 0.285714 | 19045 | 57.6 | 1 | 0 | 1 | 648 |
| 2023 | Clinton | 0.298077 | 0.274979 | 19045 | 57.6 | 1 | 0 | 1 | 648 |
| 2024 | Clinton | 0.297872 | 0.259063 | 19045 | 57.6 | 1 | 0 | 1 | 648 |
| 2017 | Crawford | 0.35619 | 0.279773 | 19047 | 55.7 | 1 | 1 | 0 | 390 |
| 2018 | Crawford | 0.357143 | 0.285978 | 19047 | 55.7 | 1 | 1 | 0 | 390 |
| 2019 | Crawford | 0.306324 | 0.225275 | 19047 | 55.7 | 1 | 1 | 0 | 390 |
| 2020 | Crawford | 0.414679 | 0.37037 | 19047 | 55.7 | 1 | 1 | 1 | 390 |
| 2021 | Crawford | 0.418079 | 0.360134 | 19047 | 55.7 | 1 | 1 | 1 | 390 |
| 2022 | Crawford | 0.408663 | 0.379019 | 19047 | 55.7 | 1 | 1 | 1 | 390 |
| 2023 | Crawford | 0.429461 | 0.410163 | 19047 | 55.7 | 1 | 1 | 1 | 390 |
| 2024 | Crawford | 0.433663 | 0.398422 | 19047 | 55.7 | 1 | 1 | 1 | 390 |
| 2017 | Dallas | 0.505504 | 0.423017 | 19049 | 69.2 | 0 | 0 | 0 | 2216 |
| 2018 | Dallas | 0.543496 | 0.50185 | 19049 | 69.2 | 0 | 0 | 0 | 2216 |
| 2019 | Dallas | 0.460232 | 0.451176 | 19049 | 69.2 | 0 | 0 | 0 | 2216 |
| 2020 | Dallas | 0.560558 | 0.545853 | 19049 | 69.2 | 0 | 0 | 1 | 2216 |
| 2021 | Dallas | 0.540206 | 0.527295 | 19049 | 69.2 | 0 | 0 | 1 | 2216 |
| 2022 | Dallas | 0.545243 | 0.542509 | 19049 | 69.2 | 0 | 0 | 1 | 2216 |
| 2023 | Dallas | 0.547156 | 0.555475 | 19049 | 69.2 | 0 | 0 | 1 | 2216 |
| 2024 | Dallas | 0.563616 | 0.553781 | 19049 | 69.2 | 0 | 0 | 1 | 2216 |
| 2017 | Davis | 0.179487 | 0.152284 | 19051 | 37.5 | 1 | 1 | 0 | 94 |
| 2018 | Davis | 0.196262 | 0.151659 | 19051 | 37.5 | 1 | 1 | 0 | 94 |
| 2019 | Davis | 0.143564 | 0.127854 | 19051 | 37.5 | 1 | 1 | 0 | 94 |
| 2020 | Davis | 0.275229 | 0.22467 | 19051 | 37.5 | 1 | 1 | 1 | 94 |
| 2021 | Davis | 0.276786 | 0.261682 | 19051 | 37.5 | 1 | 1 | 1 | 94 |
| 2022 | Davis | 0.259912 | 0.291457 | 19051 | 37.5 | 1 | 1 | 1 | 94 |
| 2023 | Davis | 0.271845 | 0.308901 | 19051 | 37.5 | 1 | 1 | 1 | 94 |
| 2024 | Davis | 0.227273 | 0.257732 | 19051 | 37.5 | 1 | 1 | 1 | 94 |
| 2017 | Decatur | 0.28 | 0.180645 | 19053 | 43.3 | 1 | 1 | 0 | 104 |
| 2018 | Decatur | 0.253165 | 0.239766 | 19053 | 43.3 | 1 | 1 | 0 | 104 |
| 2019 | Decatur | 0.251462 | 0.216867 | 19053 | 43.3 | 1 | 1 | 0 | 104 |
| 2020 | Decatur | 0.411429 | 0.336957 | 19053 | 43.3 | 1 | 1 | 1 | 104 |
| 2021 | Decatur | 0.372973 | 0.36612 | 19053 | 43.3 | 1 | 1 | 1 | 104 |
| 2022 | Decatur | 0.337079 | 0.366667 | 19053 | 43.3 | 1 | 1 | 1 | 104 |
| 2023 | Decatur | 0.319767 | 0.38125 | 19053 | 43.3 | 1 | 1 | 1 | 104 |
| 2024 | Decatur | 0.328767 | 0.304636 | 19053 | 43.3 | 1 | 1 | 1 | 104 |
| 2017 | Delaware | 0.513158 | 0.459627 | 19055 | 55.7 | 1 | 1 | 0 | 342 |
| 2018 | Delaware | 0.539936 | 0.474194 | 19055 | 55.7 | 1 | 1 | 0 | 342 |
| 2019 | Delaware | 0.449686 | 0.424242 | 19055 | 55.7 | 1 | 1 | 0 | 342 |
| 2020 | Delaware | 0.553672 | 0.530303 | 19055 | 55.7 | 1 | 1 | 1 | 342 |
| 2021 | Delaware | 0.570621 | 0.51715 | 19055 | 55.7 | 1 | 1 | 1 | 342 |
| 2022 | Delaware | 0.514451 | 0.451697 | 19055 | 55.7 | 1 | 1 | 1 | 342 |
| 2023 | Delaware | 0.506887 | 0.433526 | 19055 | 55.7 | 1 | 1 | 1 | 342 |
| 2024 | Delaware | 0.524217 | 0.447592 | 19055 | 55.7 | 1 | 1 | 1 | 342 |
| 2017 | Des Moines | 0.232481 | 0.165453 | 19057 | 53.7 | 1 | 1 | 0 | 475 |
| 2018 | Des Moines | 0.273038 | 0.208641 | 19057 | 53.7 | 1 | 1 | 0 | 475 |
| 2019 | Des Moines | 0.231335 | 0.178715 | 19057 | 53.7 | 1 | 1 | 0 | 475 |
| 2020 | Des Moines | 0.299801 | 0.231429 | 19057 | 53.7 | 1 | 1 | 1 | 475 |
| 2021 | Des Moines | 0.272727 | 0.221801 | 19057 | 53.7 | 1 | 1 | 1 | 475 |
| 2022 | Des Moines | 0.281581 | 0.235627 | 19057 | 53.7 | 1 | 1 | 1 | 475 |
| 2023 | Des Moines | 0.277666 | 0.255132 | 19057 | 53.7 | 1 | 1 | 1 | 475 |
| 2024 | Des Moines | 0.231414 | 0.221352 | 19057 | 53.7 | 1 | 1 | 1 | 475 |
| 2017 | Dickinson | 0.400612 | 0.448925 | 19059 | 62.3 | 1 | 0 | 0 | 313 |
| 2018 | Dickinson | 0.465318 | 0.474286 | 19059 | 62.3 | 1 | 0 | 0 | 313 |
| 2019 | Dickinson | 0.507205 | 0.441989 | 19059 | 62.3 | 1 | 0 | 0 | 313 |
| 2020 | Dickinson | 0.646209 | 0.588997 | 19059 | 62.3 | 1 | 0 | 1 | 313 |
| 2021 | Dickinson | 0.5 | 0.489614 | 19059 | 62.3 | 1 | 0 | 1 | 313 |
| 2022 | Dickinson | 0.503425 | 0.473846 | 19059 | 62.3 | 1 | 0 | 1 | 313 |
| 2023 | Dickinson | 0.517606 | 0.449541 | 19059 | 62.3 | 1 | 0 | 1 | 313 |
| 2024 | Dickinson | 0.450355 | 0.454545 | 19059 | 62.3 | 1 | 0 | 1 | 313 |
| 2017 | Dubuque | 0.273478 | 0.239808 | 19061 | 66.5 | 0 | 0 | 0 | 1517 |
| 2018 | Dubuque | 0.314713 | 0.286476 | 19061 | 66.5 | 0 | 0 | 0 | 1517 |
| 2019 | Dubuque | 0.271401 | 0.244573 | 19061 | 66.5 | 0 | 0 | 0 | 1517 |
| 2020 | Dubuque | 0.380527 | 0.352022 | 19061 | 66.5 | 0 | 0 | 1 | 1517 |
| 2021 | Dubuque | 0.389224 | 0.370835 | 19061 | 66.5 | 0 | 0 | 1 | 1517 |
| 2022 | Dubuque | 0.40522 | 0.372103 | 19061 | 66.5 | 0 | 0 | 1 | 1517 |
| 2023 | Dubuque | 0.425448 | 0.37244 | 19061 | 66.5 | 0 | 0 | 1 | 1517 |
| 2024 | Dubuque | 0.405601 | 0.374564 | 19061 | 66.5 | 0 | 0 | 1 | 1517 |
| 2017 | Emmet | 0.325758 | 0.285068 | 19063 | 54.3 | 1 | 1 | 0 | 162 |
| 2018 | Emmet | 0.333333 | 0.316667 | 19063 | 54.3 | 1 | 1 | 0 | 162 |
| 2019 | Emmet | 0.330508 | 0.234818 | 19063 | 54.3 | 1 | 1 | 0 | 162 |
| 2020 | Emmet | 0.403922 | 0.285171 | 19063 | 54.3 | 1 | 1 | 1 | 162 |
| 2021 | Emmet | 0.421687 | 0.27459 | 19063 | 54.3 | 1 | 1 | 1 | 162 |
| 2022 | Emmet | 0.391129 | 0.332 | 19063 | 54.3 | 1 | 1 | 1 | 162 |
| 2023 | Emmet | 0.355556 | 0.336066 | 19063 | 54.3 | 1 | 1 | 1 | 162 |
| 2024 | Emmet | 0.385366 | 0.3125 | 19063 | 54.3 | 1 | 1 | 1 | 162 |
| 2017 | Fayette | 0.490358 | 0.317708 | 19065 | 56.1 | 1 | 1 | 0 | 396 |
| 2018 | Fayette | 0.485014 | 0.43257 | 19065 | 56.1 | 1 | 1 | 0 | 396 |
| 2019 | Fayette | 0.385042 | 0.377012 | 19065 | 56.1 | 1 | 1 | 0 | 396 |
| 2020 | Fayette | 0.570352 | 0.525 | 19065 | 56.1 | 1 | 1 | 1 | 396 |
| 2021 | Fayette | 0.53886 | 0.492632 | 19065 | 56.1 | 1 | 1 | 1 | 396 |
| 2022 | Fayette | 0.535109 | 0.485777 | 19065 | 56.1 | 1 | 1 | 1 | 396 |
| 2023 | Fayette | 0.537349 | 0.468683 | 19065 | 56.1 | 1 | 1 | 1 | 396 |
| 2024 | Fayette | 0.501229 | 0.461707 | 19065 | 56.1 | 1 | 1 | 1 | 396 |
| 2017 | Floyd | 0.320132 | 0.298462 | 19067 | 55 | 1 | 1 | 0 | 238 |
| 2018 | Floyd | 0.319444 | 0.332394 | 19067 | 55 | 1 | 1 | 0 | 238 |
| 2019 | Floyd | 0.256798 | 0.277778 | 19067 | 55 | 1 | 1 | 0 | 238 |
| 2020 | Floyd | 0.404432 | 0.354286 | 19067 | 55 | 1 | 1 | 1 | 238 |
| 2021 | Floyd | 0.428962 | 0.312169 | 19067 | 55 | 1 | 1 | 1 | 238 |
| 2022 | Floyd | 0.426035 | 0.351955 | 19067 | 55 | 1 | 1 | 1 | 238 |
| 2023 | Floyd | 0.418338 | 0.318182 | 19067 | 55 | 1 | 1 | 1 | 238 |
| 2024 | Floyd | 0.410714 | 0.323529 | 19067 | 55 | 1 | 1 | 1 | 238 |
| 2017 | Franklin | 0.372449 | 0.318725 | 19069 | 55.3 | 1 | 1 | 0 | 186 |
| 2018 | Franklin | 0.406417 | 0.329545 | 19069 | 55.3 | 1 | 1 | 0 | 186 |
| 2019 | Franklin | 0.287081 | 0.294118 | 19069 | 55.3 | 1 | 1 | 0 | 186 |
| 2020 | Franklin | 0.438662 | 0.402685 | 19069 | 55.3 | 1 | 1 | 1 | 186 |
| 2021 | Franklin | 0.378906 | 0.394464 | 19069 | 55.3 | 1 | 1 | 1 | 186 |
| 2022 | Franklin | 0.396694 | 0.353333 | 19069 | 55.3 | 1 | 1 | 1 | 186 |
| 2023 | Franklin | 0.400943 | 0.373188 | 19069 | 55.3 | 1 | 1 | 1 | 186 |
| 2024 | Franklin | 0.41704 | 0.385185 | 19069 | 55.3 | 1 | 1 | 1 | 186 |
| 2017 | Fremont | 0.422535 | 0.339744 | 19071 | 54.6 | 1 | 1 | 0 | 122 |
| 2018 | Fremont | 0.409091 | 0.335443 | 19071 | 54.6 | 1 | 1 | 0 | 122 |
| 2019 | Fremont | 0.295302 | 0.3 | 19071 | 54.6 | 1 | 1 | 0 | 122 |
| 2020 | Fremont | 0.428571 | 0.405882 | 19071 | 54.6 | 1 | 1 | 1 | 122 |
| 2021 | Fremont | 0.39521 | 0.357576 | 19071 | 54.6 | 1 | 1 | 1 | 122 |
| 2022 | Fremont | 0.387097 | 0.37931 | 19071 | 54.6 | 1 | 1 | 1 | 122 |
| 2023 | Fremont | 0.331522 | 0.378049 | 19071 | 54.6 | 1 | 1 | 1 | 122 |
| 2024 | Fremont | 0.368421 | 0.381818 | 19071 | 54.6 | 1 | 1 | 1 | 122 |
| 2017 | Greene | 0.446809 | 0.42723 | 19073 | 60.3 | 1 | 0 | 0 | 192 |
| 2018 | Greene | 0.580247 | 0.447761 | 19073 | 60.3 | 1 | 0 | 0 | 192 |
| 2019 | Greene | 0.45509 | 0.389163 | 19073 | 60.3 | 1 | 0 | 0 | 192 |
| 2020 | Greene | 0.548913 | 0.520202 | 19073 | 60.3 | 1 | 0 | 1 | 192 |
| 2021 | Greene | 0.559406 | 0.513761 | 19073 | 60.3 | 1 | 0 | 1 | 192 |
| 2022 | Greene | 0.55 | 0.456731 | 19073 | 60.3 | 1 | 0 | 1 | 192 |
| 2023 | Greene | 0.530387 | 0.444954 | 19073 | 60.3 | 1 | 0 | 1 | 192 |
| 2024 | Greene | 0.555556 | 0.513228 | 19073 | 60.3 | 1 | 0 | 1 | 192 |
| 2017 | Grundy | 0.484163 | 0.384279 | 19075 | 63.6 | 0 | 0 | 0 | 294 |
| 2018 | Grundy | 0.54386 | 0.466063 | 19075 | 63.6 | 0 | 0 | 0 | 294 |
| 2019 | Grundy | 0.452991 | 0.446721 | 19075 | 63.6 | 0 | 0 | 0 | 294 |
| 2020 | Grundy | 0.601942 | 0.602787 | 19075 | 63.6 | 0 | 0 | 1 | 294 |
| 2021 | Grundy | 0.562914 | 0.623693 | 19075 | 63.6 | 0 | 0 | 1 | 294 |
| 2022 | Grundy | 0.57947 | 0.578571 | 19075 | 63.6 | 0 | 0 | 1 | 294 |
| 2023 | Grundy | 0.583051 | 0.562724 | 19075 | 63.6 | 0 | 0 | 1 | 294 |
| 2024 | Grundy | 0.623616 | 0.594502 | 19075 | 63.6 | 0 | 0 | 1 | 294 |
| 2017 | Guthrie | 0.539419 | 0.535714 | 19077 | 60.6 | 0 | 0 | 0 | 264 |
| 2018 | Guthrie | 0.577586 | 0.57197 | 19077 | 60.6 | 0 | 0 | 0 | 264 |
| 2019 | Guthrie | 0.490909 | 0.46888 | 19077 | 60.6 | 0 | 0 | 0 | 264 |
| 2020 | Guthrie | 0.593886 | 0.574899 | 19077 | 60.6 | 0 | 0 | 1 | 264 |
| 2021 | Guthrie | 0.60084 | 0.548387 | 19077 | 60.6 | 0 | 0 | 1 | 264 |
| 2022 | Guthrie | 0.576 | 0.517928 | 19077 | 60.6 | 0 | 0 | 1 | 264 |
| 2023 | Guthrie | 0.52439 | 0.528926 | 19077 | 60.6 | 0 | 0 | 1 | 264 |
| 2024 | Guthrie | 0.493724 | 0.491935 | 19077 | 60.6 | 0 | 0 | 1 | 264 |
| 2017 | Hamilton | 0.465465 | 0.335366 | 19079 | 65 | 1 | 0 | 0 | 274 |
| 2018 | Hamilton | 0.458084 | 0.34072 | 19079 | 65 | 1 | 0 | 0 | 274 |
| 2019 | Hamilton | 0.325153 | 0.25 | 19079 | 65 | 1 | 0 | 0 | 274 |
| 2020 | Hamilton | 0.450581 | 0.365432 | 19079 | 65 | 1 | 0 | 1 | 274 |
| 2021 | Hamilton | 0.432584 | 0.364611 | 19079 | 65 | 1 | 0 | 1 | 274 |
| 2022 | Hamilton | 0.458221 | 0.383754 | 19079 | 65 | 1 | 0 | 1 | 274 |
| 2023 | Hamilton | 0.447293 | 0.334311 | 19079 | 65 | 1 | 0 | 1 | 274 |
| 2024 | Hamilton | 0.449568 | 0.348974 | 19079 | 65 | 1 | 0 | 1 | 274 |
| 2017 | Hancock | 0.359375 | 0.314516 | 19081 | 51.1 | 1 | 1 | 0 | 179 |
| 2018 | Hancock | 0.409524 | 0.368852 | 19081 | 51.1 | 1 | 1 | 0 | 179 |
| 2019 | Hancock | 0.347032 | 0.309434 | 19081 | 51.1 | 1 | 1 | 0 | 179 |
| 2020 | Hancock | 0.432653 | 0.464419 | 19081 | 51.1 | 1 | 1 | 1 | 179 |
| 2021 | Hancock | 0.393939 | 0.411523 | 19081 | 51.1 | 1 | 1 | 1 | 179 |
| 2022 | Hancock | 0.367347 | 0.380282 | 19081 | 51.1 | 1 | 1 | 1 | 179 |
| 2023 | Hancock | 0.446809 | 0.37037 | 19081 | 51.1 | 1 | 1 | 1 | 179 |
| 2024 | Hancock | 0.411504 | 0.372549 | 19081 | 51.1 | 1 | 1 | 1 | 179 |
| 2017 | Hardin | 0.382813 | 0.348946 | 19083 | 57.7 | 1 | 0 | 0 | 315 |
| 2018 | Hardin | 0.411765 | 0.417661 | 19083 | 57.7 | 1 | 0 | 0 | 315 |
| 2019 | Hardin | 0.361386 | 0.345324 | 19083 | 57.7 | 1 | 0 | 0 | 315 |
| 2020 | Hardin | 0.466667 | 0.403553 | 19083 | 57.7 | 1 | 0 | 1 | 315 |
| 2021 | Hardin | 0.418367 | 0.380488 | 19083 | 57.7 | 1 | 0 | 1 | 315 |
| 2022 | Hardin | 0.407125 | 0.359897 | 19083 | 57.7 | 1 | 0 | 1 | 315 |
| 2023 | Hardin | 0.43883 | 0.360577 | 19083 | 57.7 | 1 | 0 | 1 | 315 |
| 2024 | Hardin | 0.434667 | 0.394872 | 19083 | 57.7 | 1 | 0 | 1 | 315 |
| 2017 | Harrison | 0.493789 | 0.397516 | 19085 | 56.1 | 0 | 1 | 0 | 286 |
| 2018 | Harrison | 0.478788 | 0.427653 | 19085 | 56.1 | 0 | 1 | 0 | 286 |
| 2019 | Harrison | 0.349315 | 0.314286 | 19085 | 56.1 | 0 | 1 | 0 | 286 |
| 2020 | Harrison | 0.430636 | 0.461538 | 19085 | 56.1 | 0 | 1 | 1 | 286 |
| 2021 | Harrison | 0.428571 | 0.463542 | 19085 | 56.1 | 0 | 1 | 1 | 286 |
| 2022 | Harrison | 0.408719 | 0.455497 | 19085 | 56.1 | 0 | 1 | 1 | 286 |
| 2023 | Harrison | 0.39759 | 0.4375 | 19085 | 56.1 | 0 | 1 | 1 | 286 |
| 2024 | Harrison | 0.381579 | 0.435484 | 19085 | 56.1 | 0 | 1 | 1 | 286 |
| 2017 | Henry | 0.297561 | 0.236515 | 19087 | 55.8 | 1 | 1 | 0 | 237 |
| 2018 | Henry | 0.294264 | 0.257709 | 19087 | 55.8 | 1 | 1 | 0 | 237 |
| 2019 | Henry | 0.25 | 0.211329 | 19087 | 55.8 | 1 | 1 | 0 | 237 |
| 2020 | Henry | 0.328605 | 0.286976 | 19087 | 55.8 | 1 | 1 | 1 | 237 |
| 2021 | Henry | 0.288684 | 0.279826 | 19087 | 55.8 | 1 | 1 | 1 | 237 |
| 2022 | Henry | 0.311475 | 0.247216 | 19087 | 55.8 | 1 | 1 | 1 | 237 |
| 2023 | Henry | 0.311436 | 0.254587 | 19087 | 55.8 | 1 | 1 | 1 | 237 |
| 2024 | Henry | 0.369231 | 0.284091 | 19087 | 55.8 | 1 | 1 | 1 | 237 |
| 2017 | Howard | 0.474747 | 0.366197 | 19089 | 53.8 | 1 | 1 | 0 | 193 |
| 2018 | Howard | 0.528497 | 0.474419 | 19089 | 53.8 | 1 | 1 | 0 | 193 |
| 2019 | Howard | 0.389671 | 0.425 | 19089 | 53.8 | 1 | 1 | 0 | 193 |
| 2020 | Howard | 0.497872 | 0.406504 | 19089 | 53.8 | 1 | 1 | 1 | 193 |
| 2021 | Howard | 0.434043 | 0.384937 | 19089 | 53.8 | 1 | 1 | 1 | 193 |
| 2022 | Howard | 0.434043 | 0.385892 | 19089 | 53.8 | 1 | 1 | 1 | 193 |
| 2023 | Howard | 0.454545 | 0.392694 | 19089 | 53.8 | 1 | 1 | 1 | 193 |
| 2024 | Howard | 0.478469 | 0.385321 | 19089 | 53.8 | 1 | 1 | 1 | 193 |
| 2017 | Humboldt | 0.521368 | 0.436441 | 19091 | 51.4 | 1 | 1 | 0 | 229 |
| 2018 | Humboldt | 0.543103 | 0.465217 | 19091 | 51.4 | 1 | 1 | 0 | 229 |
| 2019 | Humboldt | 0.5 | 0.457778 | 19091 | 51.4 | 1 | 1 | 0 | 229 |
| 2020 | Humboldt | 0.528384 | 0.513393 | 19091 | 51.4 | 1 | 1 | 1 | 229 |
| 2021 | Humboldt | 0.560166 | 0.497778 | 19091 | 51.4 | 1 | 1 | 1 | 229 |
| 2022 | Humboldt | 0.558036 | 0.459459 | 19091 | 51.4 | 1 | 1 | 1 | 229 |
| 2023 | Humboldt | 0.561404 | 0.481308 | 19091 | 51.4 | 1 | 1 | 1 | 229 |
| 2024 | Humboldt | 0.488038 | 0.431818 | 19091 | 51.4 | 1 | 1 | 1 | 229 |
| 2017 | Ida | 0.344262 | 0.28777 | 19093 | 50.9 | 1 | 1 | 0 | 94 |
| 2018 | Ida | 0.383333 | 0.275362 | 19093 | 50.9 | 1 | 1 | 0 | 94 |
| 2019 | Ida | 0.305556 | 0.28 | 19093 | 50.9 | 1 | 1 | 0 | 94 |
| 2020 | Ida | 0.380952 | 0.287425 | 19093 | 50.9 | 1 | 1 | 1 | 94 |
| 2021 | Ida | 0.348993 | 0.302469 | 19093 | 50.9 | 1 | 1 | 1 | 94 |
| 2022 | Ida | 0.319444 | 0.303867 | 19093 | 50.9 | 1 | 1 | 1 | 94 |
| 2023 | Ida | 0.313869 | 0.331325 | 19093 | 50.9 | 1 | 1 | 1 | 94 |
| 2024 | Ida | 0.282609 | 0.319767 | 19093 | 50.9 | 1 | 1 | 1 | 94 |
| 2017 | Iowa | 0.350769 | 0.286119 | 19095 | 64.9 | 1 | 0 | 0 | 303 |
| 2018 | Iowa | 0.4273 | 0.385714 | 19095 | 64.9 | 1 | 0 | 0 | 303 |
| 2019 | Iowa | 0.389222 | 0.355191 | 19095 | 64.9 | 1 | 0 | 0 | 303 |
| 2020 | Iowa | 0.482857 | 0.426997 | 19095 | 64.9 | 1 | 0 | 1 | 303 |
| 2021 | Iowa | 0.469565 | 0.400517 | 19095 | 64.9 | 1 | 0 | 1 | 303 |
| 2022 | Iowa | 0.460674 | 0.403101 | 19095 | 64.9 | 1 | 0 | 1 | 303 |
| 2023 | Iowa | 0.494382 | 0.465969 | 19095 | 64.9 | 1 | 0 | 1 | 303 |
| 2024 | Iowa | 0.504225 | 0.462532 | 19095 | 64.9 | 1 | 0 | 1 | 303 |
| 2017 | Jackson | 0.208075 | 0.188679 | 19097 | 54.2 | 1 | 1 | 0 | 195 |
| 2018 | Jackson | 0.25 | 0.226629 | 19097 | 54.2 | 1 | 1 | 0 | 195 |
| 2019 | Jackson | 0.21902 | 0.190981 | 19097 | 54.2 | 1 | 1 | 0 | 195 |
| 2020 | Jackson | 0.353093 | 0.253133 | 19097 | 54.2 | 1 | 1 | 1 | 195 |
| 2021 | Jackson | 0.344059 | 0.223881 | 19097 | 54.2 | 1 | 1 | 1 | 195 |
| 2022 | Jackson | 0.348958 | 0.235012 | 19097 | 54.2 | 1 | 1 | 1 | 195 |
| 2023 | Jackson | 0.335312 | 0.216019 | 19097 | 54.2 | 1 | 1 | 1 | 195 |
| 2024 | Jackson | 0.301724 | 0.23753 | 19097 | 54.2 | 1 | 1 | 1 | 195 |
| 2017 | Jasper | 0.341917 | 0.261498 | 19099 | 60.6 | 1 | 0 | 0 | 639 |
| 2018 | Jasper | 0.407913 | 0.330323 | 19099 | 60.6 | 1 | 0 | 0 | 639 |
| 2019 | Jasper | 0.380201 | 0.312581 | 19099 | 60.6 | 1 | 0 | 0 | 639 |
| 2020 | Jasper | 0.506477 | 0.444311 | 19099 | 60.6 | 1 | 0 | 1 | 639 |
| 2021 | Jasper | 0.460154 | 0.44686 | 19099 | 60.6 | 1 | 0 | 1 | 639 |
| 2022 | Jasper | 0.44793 | 0.457565 | 19099 | 60.6 | 1 | 0 | 1 | 639 |
| 2023 | Jasper | 0.442945 | 0.447205 | 19099 | 60.6 | 1 | 0 | 1 | 639 |
| 2024 | Jasper | 0.414268 | 0.437824 | 19099 | 60.6 | 1 | 0 | 1 | 639 |
| 2017 | Jefferson | 0.308333 | 0.192913 | 19101 | 50.9 | 1 | 1 | 0 | 166 |
| 2018 | Jefferson | 0.314815 | 0.23506 | 19101 | 50.9 | 1 | 1 | 0 | 166 |
| 2019 | Jefferson | 0.284698 | 0.195833 | 19101 | 50.9 | 1 | 1 | 0 | 166 |
| 2020 | Jefferson | 0.371025 | 0.327759 | 19101 | 50.9 | 1 | 1 | 1 | 166 |
| 2021 | Jefferson | 0.333333 | 0.312057 | 19101 | 50.9 | 1 | 1 | 1 | 166 |
| 2022 | Jefferson | 0.359155 | 0.291667 | 19101 | 50.9 | 1 | 1 | 1 | 166 |
| 2023 | Jefferson | 0.377622 | 0.260714 | 19101 | 50.9 | 1 | 1 | 1 | 166 |
| 2024 | Jefferson | 0.323843 | 0.310345 | 19101 | 50.9 | 1 | 1 | 1 | 166 |
| 2017 | Johnson | 0.373508 | 0.339107 | 19103 | 77.1 | 0 | 0 | 0 | 2803 |
| 2018 | Johnson | 0.490902 | 0.447407 | 19103 | 77.1 | 0 | 0 | 0 | 2803 |
| 2019 | Johnson | 0.430624 | 0.400734 | 19103 | 77.1 | 0 | 0 | 0 | 2803 |
| 2020 | Johnson | 0.520968 | 0.491115 | 19103 | 77.1 | 0 | 0 | 1 | 2803 |
| 2021 | Johnson | 0.498712 | 0.457414 | 19103 | 77.1 | 0 | 0 | 1 | 2803 |
| 2022 | Johnson | 0.492175 | 0.45505 | 19103 | 77.1 | 0 | 0 | 1 | 2803 |
| 2023 | Johnson | 0.491119 | 0.465355 | 19103 | 77.1 | 0 | 0 | 1 | 2803 |
| 2024 | Johnson | 0.487179 | 0.490087 | 19103 | 77.1 | 0 | 0 | 1 | 2803 |
| 2017 | Jones | 0.38992 | 0.30179 | 19105 | 60.5 | 0 | 0 | 0 | 412 |
| 2018 | Jones | 0.50137 | 0.418953 | 19105 | 60.5 | 0 | 0 | 0 | 412 |
| 2019 | Jones | 0.425474 | 0.381068 | 19105 | 60.5 | 0 | 0 | 0 | 412 |
| 2020 | Jones | 0.615566 | 0.573991 | 19105 | 60.5 | 0 | 0 | 1 | 412 |
| 2021 | Jones | 0.585648 | 0.537585 | 19105 | 60.5 | 0 | 0 | 1 | 412 |
| 2022 | Jones | 0.567506 | 0.5 | 19105 | 60.5 | 0 | 0 | 1 | 412 |
| 2023 | Jones | 0.53972 | 0.514019 | 19105 | 60.5 | 0 | 0 | 1 | 412 |
| 2024 | Jones | 0.556931 | 0.50939 | 19105 | 60.5 | 0 | 0 | 1 | 412 |
| 2017 | Keokuk | 0.426396 | 0.34434 | 19107 | 48.3 | 1 | 1 | 0 | 177 |
| 2018 | Keokuk | 0.455556 | 0.470588 | 19107 | 48.3 | 1 | 1 | 0 | 177 |
| 2019 | Keokuk | 0.405556 | 0.383495 | 19107 | 48.3 | 1 | 1 | 0 | 177 |
| 2020 | Keokuk | 0.481481 | 0.49505 | 19107 | 48.3 | 1 | 1 | 1 | 177 |
| 2021 | Keokuk | 0.467033 | 0.478049 | 19107 | 48.3 | 1 | 1 | 1 | 177 |
| 2022 | Keokuk | 0.507389 | 0.5 | 19107 | 48.3 | 1 | 1 | 1 | 177 |
| 2023 | Keokuk | 0.486034 | 0.415525 | 19107 | 48.3 | 1 | 1 | 1 | 177 |
| 2024 | Keokuk | 0.497409 | 0.425837 | 19107 | 48.3 | 1 | 1 | 1 | 177 |
| 2017 | Kossuth | 0.460784 | 0.365696 | 19109 | 54.1 | 1 | 1 | 0 | 315 |
| 2018 | Kossuth | 0.511182 | 0.455108 | 19109 | 54.1 | 1 | 1 | 0 | 315 |
| 2019 | Kossuth | 0.45515 | 0.41433 | 19109 | 54.1 | 1 | 1 | 0 | 315 |
| 2020 | Kossuth | 0.55942 | 0.529412 | 19109 | 54.1 | 1 | 1 | 1 | 315 |
| 2021 | Kossuth | 0.531722 | 0.496 | 19109 | 54.1 | 1 | 1 | 1 | 315 |
| 2022 | Kossuth | 0.493506 | 0.44898 | 19109 | 54.1 | 1 | 1 | 1 | 315 |
| 2023 | Kossuth | 0.44898 | 0.453552 | 19109 | 54.1 | 1 | 1 | 1 | 315 |
| 2024 | Kossuth | 0.491525 | 0.430518 | 19109 | 54.1 | 1 | 1 | 1 | 315 |
| 2017 | Lee | 0.353024 | 0.311953 | 19111 | 53.8 | 1 | 1 | 0 | 580 |
| 2018 | Lee | 0.391738 | 0.32838 | 19111 | 53.8 | 1 | 1 | 0 | 580 |
| 2019 | Lee | 0.337748 | 0.301783 | 19111 | 53.8 | 1 | 1 | 0 | 580 |
| 2020 | Lee | 0.434343 | 0.385287 | 19111 | 53.8 | 1 | 1 | 1 | 580 |
| 2021 | Lee | 0.432895 | 0.398022 | 19111 | 53.8 | 1 | 1 | 1 | 580 |
| 2022 | Lee | 0.456349 | 0.395437 | 19111 | 53.8 | 1 | 1 | 1 | 580 |
| 2023 | Lee | 0.451117 | 0.390428 | 19111 | 53.8 | 1 | 1 | 1 | 580 |
| 2024 | Lee | 0.412153 | 0.361461 | 19111 | 53.8 | 1 | 1 | 1 | 580 |
| 2017 | Linn | 0.432793 | 0.384615 | 19113 | 69.7 | 0 | 0 | 0 | 5230 |
| 2018 | Linn | 0.500927 | 0.45792 | 19113 | 69.7 | 0 | 0 | 0 | 5230 |
| 2019 | Linn | 0.453623 | 0.429077 | 19113 | 69.7 | 0 | 0 | 0 | 5230 |
| 2020 | Linn | 0.579741 | 0.539858 | 19113 | 69.7 | 0 | 0 | 1 | 5230 |
| 2021 | Linn | 0.554856 | 0.527839 | 19113 | 69.7 | 0 | 0 | 1 | 5230 |
| 2022 | Linn | 0.549792 | 0.515212 | 19113 | 69.7 | 0 | 0 | 1 | 5230 |
| 2023 | Linn | 0.545317 | 0.499082 | 19113 | 69.7 | 0 | 0 | 1 | 5230 |
| 2024 | Linn | 0.543316 | 0.506545 | 19113 | 69.7 | 0 | 0 | 1 | 5230 |
| 2017 | Louisa | 0.331797 | 0.308475 | 19115 | 54 | 1 | 1 | 0 | 191 |
| 2018 | Louisa | 0.403756 | 0.373444 | 19115 | 54 | 1 | 1 | 0 | 191 |
| 2019 | Louisa | 0.327103 | 0.335968 | 19115 | 54 | 1 | 1 | 0 | 191 |
| 2020 | Louisa | 0.488987 | 0.361217 | 19115 | 54 | 1 | 1 | 1 | 191 |
| 2021 | Louisa | 0.460905 | 0.378182 | 19115 | 54 | 1 | 1 | 1 | 191 |
| 2022 | Louisa | 0.46063 | 0.401515 | 19115 | 54 | 1 | 1 | 1 | 191 |
| 2023 | Louisa | 0.4375 | 0.34717 | 19115 | 54 | 1 | 1 | 1 | 191 |
| 2024 | Louisa | 0.391791 | 0.320313 | 19115 | 54 | 1 | 1 | 1 | 191 |
| 2017 | Lucas | 0.407407 | 0.383562 | 19117 | 46.8 | 1 | 1 | 0 | 136 |
| 2018 | Lucas | 0.407895 | 0.368098 | 19117 | 46.8 | 1 | 1 | 0 | 136 |
| 2019 | Lucas | 0.329114 | 0.274611 | 19117 | 46.8 | 1 | 1 | 0 | 136 |
| 2020 | Lucas | 0.4 | 0.372549 | 19117 | 46.8 | 1 | 1 | 1 | 136 |
| 2021 | Lucas | 0.401042 | 0.372294 | 19117 | 46.8 | 1 | 1 | 1 | 136 |
| 2022 | Lucas | 0.387435 | 0.358586 | 19117 | 46.8 | 1 | 1 | 1 | 136 |
| 2023 | Lucas | 0.404372 | 0.331633 | 19117 | 46.8 | 1 | 1 | 1 | 136 |
| 2024 | Lucas | 0.417112 | 0.328205 | 19117 | 46.8 | 1 | 1 | 1 | 136 |
| 2017 | Lyon | 0.29588 | 0.172638 | 19119 | 44.3 | 1 | 1 | 0 | 215 |
| 2018 | Lyon | 0.347682 | 0.296407 | 19119 | 44.3 | 1 | 1 | 0 | 215 |
| 2019 | Lyon | 0.289389 | 0.255193 | 19119 | 44.3 | 1 | 1 | 0 | 215 |
| 2020 | Lyon | 0.370262 | 0.352436 | 19119 | 44.3 | 1 | 1 | 1 | 215 |
| 2021 | Lyon | 0.369231 | 0.344118 | 19119 | 44.3 | 1 | 1 | 1 | 215 |
| 2022 | Lyon | 0.412903 | 0.348348 | 19119 | 44.3 | 1 | 1 | 1 | 215 |
| 2023 | Lyon | 0.415335 | 0.312312 | 19119 | 44.3 | 1 | 1 | 1 | 215 |
| 2024 | Lyon | 0.370482 | 0.35241 | 19119 | 44.3 | 1 | 1 | 1 | 215 |
| 2017 | Madison | 0.513274 | 0.453431 | 19121 | 56.3 | 0 | 0 | 0 | 407 |
| 2018 | Madison | 0.592705 | 0.5 | 19121 | 56.3 | 0 | 0 | 0 | 407 |
| 2019 | Madison | 0.542945 | 0.445313 | 19121 | 56.3 | 0 | 0 | 0 | 407 |
| 2020 | Madison | 0.575198 | 0.560411 | 19121 | 56.3 | 0 | 0 | 1 | 407 |
| 2021 | Madison | 0.591146 | 0.548781 | 19121 | 56.3 | 0 | 0 | 1 | 407 |
| 2022 | Madison | 0.580563 | 0.551546 | 19121 | 56.3 | 0 | 0 | 1 | 407 |
| 2023 | Madison | 0.532086 | 0.556373 | 19121 | 56.3 | 0 | 0 | 1 | 407 |
| 2024 | Madison | 0.507123 | 0.583333 | 19121 | 56.3 | 0 | 0 | 1 | 407 |
| 2017 | Mahaska | 0.341333 | 0.272124 | 19123 | 45.9 | 1 | 1 | 0 | 319 |
| 2018 | Mahaska | 0.359173 | 0.283517 | 19123 | 45.9 | 1 | 1 | 0 | 319 |
| 2019 | Mahaska | 0.324257 | 0.236786 | 19123 | 45.9 | 1 | 1 | 0 | 319 |
| 2020 | Mahaska | 0.424569 | 0.356604 | 19123 | 45.9 | 1 | 1 | 1 | 319 |
| 2021 | Mahaska | 0.419624 | 0.363281 | 19123 | 45.9 | 1 | 1 | 1 | 319 |
| 2022 | Mahaska | 0.389344 | 0.347913 | 19123 | 45.9 | 1 | 1 | 1 | 319 |
| 2023 | Mahaska | 0.332673 | 0.3222 | 19123 | 45.9 | 1 | 1 | 1 | 319 |
| 2024 | Mahaska | 0.327451 | 0.288499 | 19123 | 45.9 | 1 | 1 | 1 | 319 |
| 2017 | Marion | 0.358491 | 0.262402 | 19125 | 55.1 | 1 | 1 | 0 | 577 |
| 2018 | Marion | 0.41523 | 0.338275 | 19125 | 55.1 | 1 | 1 | 0 | 577 |
| 2019 | Marion | 0.341429 | 0.28534 | 19125 | 55.1 | 1 | 1 | 0 | 577 |
| 2020 | Marion | 0.455381 | 0.396938 | 19125 | 55.1 | 1 | 1 | 1 | 577 |
| 2021 | Marion | 0.422611 | 0.413911 | 19125 | 55.1 | 1 | 1 | 1 | 577 |
| 2022 | Marion | 0.413929 | 0.395566 | 19125 | 55.1 | 1 | 1 | 1 | 577 |
| 2023 | Marion | 0.397436 | 0.342007 | 19125 | 55.1 | 1 | 1 | 1 | 577 |
| 2024 | Marion | 0.389447 | 0.330739 | 19125 | 55.1 | 1 | 1 | 1 | 577 |
| 2017 | Marshall | 0.576471 | 0.505088 | 19127 | 68.4 | 1 | 0 | 0 | 1248 |
| 2018 | Marshall | 0.563953 | 0.545374 | 19127 | 68.4 | 1 | 0 | 0 | 1248 |
| 2019 | Marshall | 0.506641 | 0.505163 | 19127 | 68.4 | 1 | 0 | 0 | 1248 |
| 2020 | Marshall | 0.587601 | 0.576132 | 19127 | 68.4 | 1 | 0 | 1 | 1248 |
| 2021 | Marshall | 0.58165 | 0.524411 | 19127 | 68.4 | 1 | 0 | 1 | 1248 |
| 2022 | Marshall | 0.58981 | 0.539474 | 19127 | 68.4 | 1 | 0 | 1 | 1248 |
| 2023 | Marshall | 0.570925 | 0.52618 | 19127 | 68.4 | 1 | 0 | 1 | 1248 |
| 2024 | Marshall | 0.568431 | 0.547725 | 19127 | 68.4 | 1 | 0 | 1 | 1248 |
| 2017 | Mills | 0.320755 | 0.29558 | 19129 | 58 | 0 | 0 | 0 | 236 |
| 2018 | Mills | 0.37224 | 0.322857 | 19129 | 58 | 0 | 0 | 0 | 236 |
| 2019 | Mills | 0.337621 | 0.267241 | 19129 | 58 | 0 | 0 | 0 | 236 |
| 2020 | Mills | 0.363095 | 0.353403 | 19129 | 58 | 0 | 0 | 1 | 236 |
| 2021 | Mills | 0.336158 | 0.302326 | 19129 | 58 | 0 | 0 | 1 | 236 |
| 2022 | Mills | 0.357527 | 0.32973 | 19129 | 58 | 0 | 0 | 1 | 236 |
| 2023 | Mills | 0.348958 | 0.314208 | 19129 | 58 | 0 | 0 | 1 | 236 |
| 2024 | Mills | 0.356948 | 0.332353 | 19129 | 58 | 0 | 0 | 1 | 236 |
| 2017 | Mitchell | 0.491803 | 0.349727 | 19131 | 49.9 | 1 | 1 | 0 | 181 |
| 2018 | Mitchell | 0.446809 | 0.46114 | 19131 | 49.9 | 1 | 1 | 0 | 181 |
| 2019 | Mitchell | 0.333333 | 0.402116 | 19131 | 49.9 | 1 | 1 | 0 | 181 |
| 2020 | Mitchell | 0.517413 | 0.453704 | 19131 | 49.9 | 1 | 1 | 1 | 181 |
| 2021 | Mitchell | 0.450237 | 0.447368 | 19131 | 49.9 | 1 | 1 | 1 | 181 |
| 2022 | Mitchell | 0.463964 | 0.432558 | 19131 | 49.9 | 1 | 1 | 1 | 181 |
| 2023 | Mitchell | 0.434959 | 0.424779 | 19131 | 49.9 | 1 | 1 | 1 | 181 |
| 2024 | Mitchell | 0.466667 | 0.4 | 19131 | 49.9 | 1 | 1 | 1 | 181 |
| 2017 | Monona | 0.551887 | 0.475728 | 19133 | 55.4 | 1 | 1 | 0 | 207 |
| 2018 | Monona | 0.595122 | 0.536842 | 19133 | 55.4 | 1 | 1 | 0 | 207 |
| 2019 | Monona | 0.561576 | 0.514706 | 19133 | 55.4 | 1 | 1 | 0 | 207 |
| 2020 | Monona | 0.570732 | 0.518349 | 19133 | 55.4 | 1 | 1 | 1 | 207 |
| 2021 | Monona | 0.557214 | 0.525581 | 19133 | 55.4 | 1 | 1 | 1 | 207 |
| 2022 | Monona | 0.536585 | 0.452632 | 19133 | 55.4 | 1 | 1 | 1 | 207 |
| 2023 | Monona | 0.502463 | 0.463687 | 19133 | 55.4 | 1 | 1 | 1 | 207 |
| 2024 | Monona | 0.478947 | 0.422619 | 19133 | 55.4 | 1 | 1 | 1 | 207 |
| 2017 | Monroe | 0.428571 | 0.2875 | 19135 | 47.7 | 1 | 1 | 0 | 121 |
| 2018 | Monroe | 0.43125 | 0.397436 | 19135 | 47.7 | 1 | 1 | 0 | 121 |
| 2019 | Monroe | 0.278788 | 0.288462 | 19135 | 47.7 | 1 | 1 | 0 | 121 |
| 2020 | Monroe | 0.394737 | 0.376884 | 19135 | 47.7 | 1 | 1 | 1 | 121 |
| 2021 | Monroe | 0.434783 | 0.335 | 19135 | 47.7 | 1 | 1 | 1 | 121 |
| 2022 | Monroe | 0.422619 | 0.273196 | 19135 | 47.7 | 1 | 1 | 1 | 121 |
| 2023 | Monroe | 0.355422 | 0.271676 | 19135 | 47.7 | 1 | 1 | 1 | 121 |
| 2024 | Monroe | 0.313609 | 0.291391 | 19135 | 47.7 | 1 | 1 | 1 | 121 |
| 2017 | Montgomery | 0.387097 | 0.270492 | 19137 | 59.6 | 1 | 0 | 0 | 189 |
| 2018 | Montgomery | 0.410377 | 0.2827 | 19137 | 59.6 | 1 | 0 | 0 | 189 |
| 2019 | Montgomery | 0.365297 | 0.320313 | 19137 | 59.6 | 1 | 0 | 0 | 189 |
| 2020 | Montgomery | 0.46473 | 0.3861 | 19137 | 59.6 | 1 | 0 | 1 | 189 |
| 2021 | Montgomery | 0.423611 | 0.380392 | 19137 | 59.6 | 1 | 0 | 1 | 189 |
| 2022 | Montgomery | 0.455197 | 0.446721 | 19137 | 59.6 | 1 | 0 | 1 | 189 |
| 2023 | Montgomery | 0.413934 | 0.396624 | 19137 | 59.6 | 1 | 0 | 1 | 189 |
| 2024 | Montgomery | 0.412844 | 0.398305 | 19137 | 59.6 | 1 | 0 | 1 | 189 |
| 2017 | Muscatine | 0.406433 | 0.315978 | 19139 | 63.8 | 1 | 0 | 0 | 909 |
| 2018 | Muscatine | 0.447552 | 0.381853 | 19139 | 63.8 | 1 | 0 | 0 | 909 |
| 2019 | Muscatine | 0.388676 | 0.355472 | 19139 | 63.8 | 1 | 0 | 0 | 909 |
| 2020 | Muscatine | 0.470211 | 0.444341 | 19139 | 63.8 | 1 | 0 | 1 | 909 |
| 2021 | Muscatine | 0.44007 | 0.412489 | 19139 | 63.8 | 1 | 0 | 1 | 909 |
| 2022 | Muscatine | 0.451082 | 0.393327 | 19139 | 63.8 | 1 | 0 | 1 | 909 |
| 2023 | Muscatine | 0.45479 | 0.412073 | 19139 | 63.8 | 1 | 0 | 1 | 909 |
| 2024 | Muscatine | 0.463985 | 0.417593 | 19139 | 63.8 | 1 | 0 | 1 | 909 |
| 2017 | OBrien | 0.436893 | 0.427441 | 19141 | 52.1 | 1 | 1 | 0 | 323 |
| 2018 | OBrien | 0.509554 | 0.427461 | 19141 | 52.1 | 1 | 1 | 0 | 323 |
| 2019 | OBrien | 0.460064 | 0.378667 | 19141 | 52.1 | 1 | 1 | 0 | 323 |
| 2020 | OBrien | 0.498433 | 0.498708 | 19141 | 52.1 | 1 | 1 | 1 | 323 |
| 2021 | OBrien | 0.498452 | 0.464286 | 19141 | 52.1 | 1 | 1 | 1 | 323 |
| 2022 | OBrien | 0.5 | 0.482143 | 19141 | 52.1 | 1 | 1 | 1 | 323 |
| 2023 | OBrien | 0.535294 | 0.457478 | 19141 | 52.1 | 1 | 1 | 1 | 323 |
| 2024 | OBrien | 0.519757 | 0.462908 | 19141 | 52.1 | 1 | 1 | 1 | 323 |
| 2017 | Osceola | 0.376 | 0.34375 | 19143 | 46.6 | 1 | 1 | 0 | 121 |
| 2018 | Osceola | 0.364964 | 0.396226 | 19143 | 46.6 | 1 | 1 | 0 | 121 |
| 2019 | Osceola | 0.314685 | 0.372549 | 19143 | 46.6 | 1 | 1 | 0 | 121 |
| 2020 | Osceola | 0.471015 | 0.493243 | 19143 | 46.6 | 1 | 1 | 1 | 121 |
| 2021 | Osceola | 0.492537 | 0.522727 | 19143 | 46.6 | 1 | 1 | 1 | 121 |
| 2022 | Osceola | 0.5 | 0.534351 | 19143 | 46.6 | 1 | 1 | 1 | 121 |
| 2023 | Osceola | 0.439716 | 0.40625 | 19143 | 46.6 | 1 | 1 | 1 | 121 |
| 2024 | Osceola | 0.507692 | 0.366906 | 19143 | 46.6 | 1 | 1 | 1 | 121 |
| 2017 | Page | 0.402899 | 0.396325 | 19145 | 56.3 | 1 | 0 | 0 | 346 |
| 2018 | Page | 0.450704 | 0.476316 | 19145 | 56.3 | 1 | 0 | 0 | 346 |
| 2019 | Page | 0.424242 | 0.417085 | 19145 | 56.3 | 1 | 0 | 0 | 346 |
| 2020 | Page | 0.5 | 0.508393 | 19145 | 56.3 | 1 | 0 | 1 | 346 |
| 2021 | Page | 0.506887 | 0.498753 | 19145 | 56.3 | 1 | 0 | 1 | 346 |
| 2022 | Page | 0.507463 | 0.468421 | 19145 | 56.3 | 1 | 0 | 1 | 346 |
| 2023 | Page | 0.494819 | 0.456284 | 19145 | 56.3 | 1 | 0 | 1 | 346 |
| 2024 | Page | 0.49435 | 0.461126 | 19145 | 56.3 | 1 | 0 | 1 | 346 |
| 2017 | Palo Alto | 0.361386 | 0.327189 | 19147 | 52.4 | 1 | 1 | 0 | 144 |
| 2018 | Palo Alto | 0.44 | 0.366492 | 19147 | 52.4 | 1 | 1 | 0 | 144 |
| 2019 | Palo Alto | 0.350515 | 0.304762 | 19147 | 52.4 | 1 | 1 | 0 | 144 |
| 2020 | Palo Alto | 0.467337 | 0.402985 | 19147 | 52.4 | 1 | 1 | 1 | 144 |
| 2021 | Palo Alto | 0.39 | 0.349754 | 19147 | 52.4 | 1 | 1 | 1 | 144 |
| 2022 | Palo Alto | 0.350254 | 0.347368 | 19147 | 52.4 | 1 | 1 | 1 | 144 |
| 2023 | Palo Alto | 0.407407 | 0.343915 | 19147 | 52.4 | 1 | 1 | 1 | 144 |
| 2024 | Palo Alto | 0.40678 | 0.292135 | 19147 | 52.4 | 1 | 1 | 1 | 144 |
| 2017 | Plymouth | 0.402135 | 0.288136 | 19149 | 53.8 | 0 | 1 | 0 | 540 |
| 2018 | Plymouth | 0.461947 | 0.373239 | 19149 | 53.8 | 0 | 1 | 0 | 540 |
| 2019 | Plymouth | 0.402116 | 0.365 | 19149 | 53.8 | 0 | 1 | 0 | 540 |
| 2020 | Plymouth | 0.500775 | 0.508449 | 19149 | 53.8 | 0 | 1 | 1 | 540 |
| 2021 | Plymouth | 0.479688 | 0.502152 | 19149 | 53.8 | 0 | 1 | 1 | 540 |
| 2022 | Plymouth | 0.469419 | 0.467532 | 19149 | 53.8 | 0 | 1 | 1 | 540 |
| 2023 | Plymouth | 0.438017 | 0.408889 | 19149 | 53.8 | 0 | 1 | 1 | 540 |
| 2024 | Plymouth | 0.440735 | 0.385307 | 19149 | 53.8 | 0 | 1 | 1 | 540 |
| 2017 | Pocahontas | 0.47482 | 0.321678 | 19151 | 54.4 | 1 | 1 | 0 | 138 |
| 2018 | Pocahontas | 0.517483 | 0.445946 | 19151 | 54.4 | 1 | 1 | 0 | 138 |
| 2019 | Pocahontas | 0.430657 | 0.372549 | 19151 | 54.4 | 1 | 1 | 0 | 138 |
| 2020 | Pocahontas | 0.522876 | 0.470968 | 19151 | 54.4 | 1 | 1 | 1 | 138 |
| 2021 | Pocahontas | 0.493056 | 0.443038 | 19151 | 54.4 | 1 | 1 | 1 | 138 |
| 2022 | Pocahontas | 0.506494 | 0.506494 | 19151 | 54.4 | 1 | 1 | 1 | 138 |
| 2023 | Pocahontas | 0.533784 | 0.510345 | 19151 | 54.4 | 1 | 1 | 1 | 138 |
| 2024 | Pocahontas | 0.464052 | 0.5 | 19151 | 54.4 | 1 | 1 | 1 | 138 |
| 2017 | Polk | 0.458997 | 0.398345 | 19153 | 71.2 | 0 | 0 | 0 | 12351 |
| 2018 | Polk | 0.505093 | 0.45508 | 19153 | 71.2 | 0 | 0 | 0 | 12351 |
| 2019 | Polk | 0.448725 | 0.414097 | 19153 | 71.2 | 0 | 0 | 0 | 12351 |
| 2020 | Polk | 0.545619 | 0.522202 | 19153 | 71.2 | 0 | 0 | 1 | 12351 |
| 2021 | Polk | 0.527502 | 0.515615 | 19153 | 71.2 | 0 | 0 | 1 | 12351 |
| 2022 | Polk | 0.532103 | 0.50686 | 19153 | 71.2 | 0 | 0 | 1 | 12351 |
| 2023 | Polk | 0.533216 | 0.501501 | 19153 | 71.2 | 0 | 0 | 1 | 12351 |
| 2024 | Polk | 0.52985 | 0.498152 | 19153 | 71.2 | 0 | 0 | 1 | 12351 |
| 2017 | Pottawattamie | 0.421154 | 0.371543 | 19155 | 62.1 | 0 | 0 | 0 | 1900 |
| 2018 | Pottawattamie | 0.433333 | 0.395133 | 19155 | 62.1 | 0 | 0 | 0 | 1900 |
| 2019 | Pottawattamie | 0.37196 | 0.348592 | 19155 | 62.1 | 0 | 0 | 0 | 1900 |
| 2020 | Pottawattamie | 0.437418 | 0.427377 | 19155 | 62.1 | 0 | 0 | 1 | 1900 |
| 2021 | Pottawattamie | 0.428207 | 0.428205 | 19155 | 62.1 | 0 | 0 | 1 | 1900 |
| 2022 | Pottawattamie | 0.42798 | 0.427151 | 19155 | 62.1 | 0 | 0 | 1 | 1900 |
| 2023 | Pottawattamie | 0.451505 | 0.440224 | 19155 | 62.1 | 0 | 0 | 1 | 1900 |
| 2024 | Pottawattamie | 0.441253 | 0.420348 | 19155 | 62.1 | 0 | 0 | 1 | 1900 |
| 2017 | Poweshiek | 0.518135 | 0.478947 | 19157 | 63 | 1 | 0 | 0 | 377 |
| 2018 | Poweshiek | 0.538259 | 0.542105 | 19157 | 63 | 1 | 0 | 0 | 377 |
| 2019 | Poweshiek | 0.481579 | 0.469496 | 19157 | 63 | 1 | 0 | 0 | 377 |
| 2020 | Poweshiek | 0.554645 | 0.498701 | 19157 | 63 | 1 | 0 | 1 | 377 |
| 2021 | Poweshiek | 0.518135 | 0.488491 | 19157 | 63 | 1 | 0 | 1 | 377 |
| 2022 | Poweshiek | 0.467192 | 0.454768 | 19157 | 63 | 1 | 0 | 1 | 377 |
| 2023 | Poweshiek | 0.451852 | 0.45 | 19157 | 63 | 1 | 0 | 1 | 377 |
| 2024 | Poweshiek | 0.440299 | 0.451444 | 19157 | 63 | 1 | 0 | 1 | 377 |
| 2017 | Ringgold | 0.49505 | 0.430108 | 19159 | 54.1 | 1 | 1 | 0 | 104 |
| 2018 | Ringgold | 0.572917 | 0.447368 | 19159 | 54.1 | 1 | 1 | 0 | 104 |
| 2019 | Ringgold | 0.465116 | 0.420561 | 19159 | 54.1 | 1 | 1 | 0 | 104 |
| 2020 | Ringgold | 0.481818 | 0.452381 | 19159 | 54.1 | 1 | 1 | 1 | 104 |
| 2021 | Ringgold | 0.551402 | 0.458716 | 19159 | 54.1 | 1 | 1 | 1 | 104 |
| 2022 | Ringgold | 0.508475 | 0.495652 | 19159 | 54.1 | 1 | 1 | 1 | 104 |
| 2023 | Ringgold | 0.509434 | 0.509091 | 19159 | 54.1 | 1 | 1 | 1 | 104 |
| 2024 | Ringgold | 0.639535 | 0.574713 | 19159 | 54.1 | 1 | 1 | 1 | 104 |
| 2017 | Sac | 0.392857 | 0.304348 | 19161 | 54.3 | 1 | 1 | 0 | 157 |
| 2018 | Sac | 0.419355 | 0.335052 | 19161 | 54.3 | 1 | 1 | 0 | 157 |
| 2019 | Sac | 0.286996 | 0.248804 | 19161 | 54.3 | 1 | 1 | 0 | 157 |
| 2020 | Sac | 0.471111 | 0.410714 | 19161 | 54.3 | 1 | 1 | 1 | 157 |
| 2021 | Sac | 0.4375 | 0.38756 | 19161 | 54.3 | 1 | 1 | 1 | 157 |
| 2022 | Sac | 0.447489 | 0.375 | 19161 | 54.3 | 1 | 1 | 1 | 157 |
| 2023 | Sac | 0.460396 | 0.330097 | 19161 | 54.3 | 1 | 1 | 1 | 157 |
| 2024 | Sac | 0.44382 | 0.306533 | 19161 | 54.3 | 1 | 1 | 1 | 157 |
| 2017 | Scott | 0.382969 | 0.336595 | 19163 | 66.9 | 0 | 0 | 0 | 3444 |
| 2018 | Scott | 0.397678 | 0.376095 | 19163 | 66.9 | 0 | 0 | 0 | 3444 |
| 2019 | Scott | 0.356204 | 0.33245 | 19163 | 66.9 | 0 | 0 | 0 | 3444 |
| 2020 | Scott | 0.421404 | 0.391142 | 19163 | 66.9 | 0 | 0 | 1 | 3444 |
| 2021 | Scott | 0.395811 | 0.364823 | 19163 | 66.9 | 0 | 0 | 1 | 3444 |
| 2022 | Scott | 0.392663 | 0.354282 | 19163 | 66.9 | 0 | 0 | 1 | 3444 |
| 2023 | Scott | 0.3783 | 0.36017 | 19163 | 66.9 | 0 | 0 | 1 | 3444 |
| 2024 | Scott | 0.376123 | 0.359491 | 19163 | 66.9 | 0 | 0 | 1 | 3444 |
| 2017 | Shelby | 0.470817 | 0.492701 | 19165 | 61.3 | 1 | 0 | 0 | 236 |
| 2018 | Shelby | 0.415254 | 0.47541 | 19165 | 61.3 | 1 | 0 | 0 | 236 |
| 2019 | Shelby | 0.372951 | 0.404167 | 19165 | 61.3 | 1 | 0 | 0 | 236 |
| 2020 | Shelby | 0.489726 | 0.46831 | 19165 | 61.3 | 1 | 0 | 1 | 236 |
| 2021 | Shelby | 0.498208 | 0.469799 | 19165 | 61.3 | 1 | 0 | 1 | 236 |
| 2022 | Shelby | 0.476563 | 0.435374 | 19165 | 61.3 | 1 | 0 | 1 | 236 |
| 2023 | Shelby | 0.443478 | 0.434615 | 19165 | 61.3 | 1 | 0 | 1 | 236 |
| 2024 | Shelby | 0.445946 | 0.441948 | 19165 | 61.3 | 1 | 0 | 1 | 236 |
| 2017 | Sioux | 0.341703 | 0.317597 | 19167 | 43.2 | 1 | 1 | 0 | 723 |
| 2018 | Sioux | 0.35383 | 0.341262 | 19167 | 43.2 | 1 | 1 | 0 | 723 |
| 2019 | Sioux | 0.308943 | 0.278008 | 19167 | 43.2 | 1 | 1 | 0 | 723 |
| 2020 | Sioux | 0.44873 | 0.352262 | 19167 | 43.2 | 1 | 1 | 1 | 723 |
| 2021 | Sioux | 0.41791 | 0.347249 | 19167 | 43.2 | 1 | 1 | 1 | 723 |
| 2022 | Sioux | 0.399809 | 0.350781 | 19167 | 43.2 | 1 | 1 | 1 | 723 |
| 2023 | Sioux | 0.364349 | 0.327206 | 19167 | 43.2 | 1 | 1 | 1 | 723 |
| 2024 | Sioux | 0.357422 | 0.34903 | 19167 | 43.2 | 1 | 1 | 1 | 723 |
| 2017 | Story | 0.457638 | 0.415894 | 19169 | 66 | 0 | 0 | 0 | 1585 |
| 2018 | Story | 0.492136 | 0.471529 | 19169 | 66 | 0 | 0 | 0 | 1585 |
| 2019 | Story | 0.426821 | 0.407063 | 19169 | 66 | 0 | 0 | 0 | 1585 |
| 2020 | Story | 0.513577 | 0.508159 | 19169 | 66 | 0 | 0 | 1 | 1585 |
| 2021 | Story | 0.483653 | 0.491413 | 19169 | 66 | 0 | 0 | 1 | 1585 |
| 2022 | Story | 0.466298 | 0.466238 | 19169 | 66 | 0 | 0 | 1 | 1585 |
| 2023 | Story | 0.456654 | 0.442544 | 19169 | 66 | 0 | 0 | 1 | 1585 |
| 2024 | Story | 0.465423 | 0.440232 | 19169 | 66 | 0 | 0 | 1 | 1585 |
| 2017 | Tama | 0.475703 | 0.50625 | 19171 | 64.7 | 1 | 0 | 0 | 458 |
| 2018 | Tama | 0.539216 | 0.526539 | 19171 | 64.7 | 1 | 0 | 0 | 458 |
| 2019 | Tama | 0.47561 | 0.448052 | 19171 | 64.7 | 1 | 0 | 0 | 458 |
| 2020 | Tama | 0.580336 | 0.563636 | 19171 | 64.7 | 1 | 0 | 1 | 458 |
| 2021 | Tama | 0.538813 | 0.524027 | 19171 | 64.7 | 1 | 0 | 1 | 458 |
| 2022 | Tama | 0.569476 | 0.559494 | 19171 | 64.7 | 1 | 0 | 1 | 458 |
| 2023 | Tama | 0.574944 | 0.553571 | 19171 | 64.7 | 1 | 0 | 1 | 458 |
| 2024 | Tama | 0.612293 | 0.543536 | 19171 | 64.7 | 1 | 0 | 1 | 458 |
| 2017 | Taylor | 0.485714 | 0.466667 | 19173 | 48.6 | 1 | 1 | 0 | 129 |
| 2018 | Taylor | 0.524138 | 0.440252 | 19173 | 48.6 | 1 | 1 | 0 | 129 |
| 2019 | Taylor | 0.379518 | 0.369942 | 19173 | 48.6 | 1 | 1 | 0 | 129 |
| 2020 | Taylor | 0.546012 | 0.493506 | 19173 | 48.6 | 1 | 1 | 1 | 129 |
| 2021 | Taylor | 0.453333 | 0.483871 | 19173 | 48.6 | 1 | 1 | 1 | 129 |
| 2022 | Taylor | 0.448 | 0.432836 | 19173 | 48.6 | 1 | 1 | 1 | 129 |
| 2023 | Taylor | 0.462963 | 0.482143 | 19173 | 48.6 | 1 | 1 | 1 | 129 |
| 2024 | Taylor | 0.402174 | 0.496 | 19173 | 48.6 | 1 | 1 | 1 | 129 |
| 2017 | Union | 0.518797 | 0.400697 | 19175 | 53.6 | 1 | 1 | 0 | 281 |
| 2018 | Union | 0.561983 | 0.509294 | 19175 | 53.6 | 1 | 1 | 0 | 281 |
| 2019 | Union | 0.463602 | 0.39172 | 19175 | 53.6 | 1 | 1 | 0 | 281 |
| 2020 | Union | 0.560284 | 0.498525 | 19175 | 53.6 | 1 | 1 | 1 | 281 |
| 2021 | Union | 0.496429 | 0.453757 | 19175 | 53.6 | 1 | 1 | 1 | 281 |
| 2022 | Union | 0.57377 | 0.547101 | 19175 | 53.6 | 1 | 1 | 1 | 281 |
| 2023 | Union | 0.590909 | 0.557692 | 19175 | 53.6 | 1 | 1 | 1 | 281 |
| 2024 | Union | 0.560606 | 0.496063 | 19175 | 53.6 | 1 | 1 | 1 | 281 |
| 2017 | Van Buren | 0.251799 | 0.210526 | 19177 | 43.9 | 1 | 1 | 0 | 72 |
| 2018 | Van Buren | 0.277778 | 0.261146 | 19177 | 43.9 | 1 | 1 | 0 | 72 |
| 2019 | Van Buren | 0.238095 | 0.239766 | 19177 | 43.9 | 1 | 1 | 0 | 72 |
| 2020 | Van Buren | 0.372093 | 0.325301 | 19177 | 43.9 | 1 | 1 | 1 | 72 |
| 2021 | Van Buren | 0.315385 | 0.22619 | 19177 | 43.9 | 1 | 1 | 1 | 72 |
| 2022 | Van Buren | 0.207407 | 0.2125 | 19177 | 43.9 | 1 | 1 | 1 | 72 |
| 2023 | Van Buren | 0.22963 | 0.196319 | 19177 | 43.9 | 1 | 1 | 1 | 72 |
| 2024 | Van Buren | 0.235714 | 0.177914 | 19177 | 43.9 | 1 | 1 | 1 | 72 |
| 2017 | Wapello | 0.390273 | 0.32526 | 19179 | 51.4 | 1 | 1 | 0 | 735 |
| 2018 | Wapello | 0.397041 | 0.382253 | 19179 | 51.4 | 1 | 1 | 0 | 735 |
| 2019 | Wapello | 0.372842 | 0.348023 | 19179 | 51.4 | 1 | 1 | 0 | 735 |
| 2020 | Wapello | 0.446203 | 0.42623 | 19179 | 51.4 | 1 | 1 | 1 | 735 |
| 2021 | Wapello | 0.448677 | 0.38668 | 19179 | 51.4 | 1 | 1 | 1 | 735 |
| 2022 | Wapello | 0.421218 | 0.393281 | 19179 | 51.4 | 1 | 1 | 1 | 735 |
| 2023 | Wapello | 0.416667 | 0.38191 | 19179 | 51.4 | 1 | 1 | 1 | 735 |
| 2024 | Wapello | 0.423374 | 0.382805 | 19179 | 51.4 | 1 | 1 | 1 | 735 |
| 2017 | Warren | 0.460159 | 0.390668 | 19181 | 62.6 | 0 | 0 | 0 | 1235 |
| 2018 | Warren | 0.50414 | 0.451473 | 19181 | 62.6 | 0 | 0 | 0 | 1235 |
| 2019 | Warren | 0.421373 | 0.38188 | 19181 | 62.6 | 0 | 0 | 0 | 1235 |
| 2020 | Warren | 0.561376 | 0.54973 | 19181 | 62.6 | 0 | 0 | 1 | 1235 |
| 2021 | Warren | 0.550473 | 0.528081 | 19181 | 62.6 | 0 | 0 | 1 | 1235 |
| 2022 | Warren | 0.5656 | 0.504399 | 19181 | 62.6 | 0 | 0 | 1 | 1235 |
| 2023 | Warren | 0.571313 | 0.509158 | 19181 | 62.6 | 0 | 0 | 1 | 1235 |
| 2024 | Warren | 0.555644 | 0.504913 | 19181 | 62.6 | 0 | 0 | 1 | 1235 |
| 2017 | Washington | 0.354379 | 0.317757 | 19183 | 59.1 | 0 | 0 | 0 | 400 |
| 2018 | Washington | 0.409978 | 0.356089 | 19183 | 59.1 | 0 | 0 | 0 | 400 |
| 2019 | Washington | 0.343035 | 0.319846 | 19183 | 59.1 | 0 | 0 | 0 | 400 |
| 2020 | Washington | 0.465447 | 0.44358 | 19183 | 59.1 | 0 | 0 | 1 | 400 |
| 2021 | Washington | 0.443378 | 0.422265 | 19183 | 59.1 | 0 | 0 | 1 | 400 |
| 2022 | Washington | 0.424419 | 0.411306 | 19183 | 59.1 | 0 | 0 | 1 | 400 |
| 2023 | Washington | 0.42094 | 0.403475 | 19183 | 59.1 | 0 | 0 | 1 | 400 |
| 2024 | Washington | 0.436681 | 0.400402 | 19183 | 59.1 | 0 | 0 | 1 | 400 |
| 2017 | Wayne | 0.384615 | 0.308271 | 19185 | 43.5 | 1 | 1 | 0 | 99 |
| 2018 | Wayne | 0.4 | 0.37037 | 19185 | 43.5 | 1 | 1 | 0 | 99 |
| 2019 | Wayne | 0.349593 | 0.333333 | 19185 | 43.5 | 1 | 1 | 0 | 99 |
| 2020 | Wayne | 0.444444 | 0.3125 | 19185 | 43.5 | 1 | 1 | 1 | 99 |
| 2021 | Wayne | 0.394366 | 0.30137 | 19185 | 43.5 | 1 | 1 | 1 | 99 |
| 2022 | Wayne | 0.388889 | 0.343949 | 19185 | 43.5 | 1 | 1 | 1 | 99 |
| 2023 | Wayne | 0.37594 | 0.314815 | 19185 | 43.5 | 1 | 1 | 1 | 99 |
| 2024 | Wayne | 0.352941 | 0.297468 | 19185 | 43.5 | 1 | 1 | 1 | 99 |
| 2017 | Webster | 0.417462 | 0.307951 | 19187 | 60 | 1 | 0 | 0 | 692 |
| 2018 | Webster | 0.42973 | 0.375729 | 19187 | 60 | 1 | 0 | 0 | 692 |
| 2019 | Webster | 0.374841 | 0.331281 | 19187 | 60 | 1 | 0 | 0 | 692 |
| 2020 | Webster | 0.482412 | 0.441527 | 19187 | 60 | 1 | 0 | 1 | 692 |
| 2021 | Webster | 0.489924 | 0.433451 | 19187 | 60 | 1 | 0 | 1 | 692 |
| 2022 | Webster | 0.493557 | 0.439394 | 19187 | 60 | 1 | 0 | 1 | 692 |
| 2023 | Webster | 0.497396 | 0.423358 | 19187 | 60 | 1 | 0 | 1 | 692 |
| 2024 | Webster | 0.494253 | 0.457921 | 19187 | 60 | 1 | 0 | 1 | 692 |
| 2017 | Winnebago | 0.261803 | 0.265918 | 19189 | 54.7 | 1 | 1 | 0 | 158 |
| 2018 | Winnebago | 0.371795 | 0.30292 | 19189 | 54.7 | 1 | 1 | 0 | 158 |
| 2019 | Winnebago | 0.294118 | 0.296578 | 19189 | 54.7 | 1 | 1 | 0 | 158 |
| 2020 | Winnebago | 0.344262 | 0.342975 | 19189 | 54.7 | 1 | 1 | 1 | 158 |
| 2021 | Winnebago | 0.334601 | 0.336134 | 19189 | 54.7 | 1 | 1 | 1 | 158 |
| 2022 | Winnebago | 0.346939 | 0.294643 | 19189 | 54.7 | 1 | 1 | 1 | 158 |
| 2023 | Winnebago | 0.37751 | 0.30531 | 19189 | 54.7 | 1 | 1 | 1 | 158 |
| 2024 | Winnebago | 0.390244 | 0.288889 | 19189 | 54.7 | 1 | 1 | 1 | 158 |
| 2017 | Winneshiek | 0.6 | 0.487981 | 19191 | 65 | 1 | 0 | 0 | 423 |
| 2018 | Winneshiek | 0.549865 | 0.520408 | 19191 | 65 | 1 | 0 | 0 | 423 |
| 2019 | Winneshiek | 0.458015 | 0.435356 | 19191 | 65 | 1 | 0 | 0 | 423 |
| 2020 | Winneshiek | 0.583333 | 0.540984 | 19191 | 65 | 1 | 0 | 1 | 423 |
| 2021 | Winneshiek | 0.568965 | 0.546318 | 19191 | 65 | 1 | 0 | 1 | 423 |
| 2022 | Winneshiek | 0.613158 | 0.546763 | 19191 | 65 | 1 | 0 | 1 | 423 |
| 2023 | Winneshiek | 0.606607 | 0.563342 | 19191 | 65 | 1 | 0 | 1 | 423 |
| 2024 | Winneshiek | 0.601852 | 0.611268 | 19191 | 65 | 1 | 0 | 1 | 423 |
| 2017 | Woodbury | 0.380407 | 0.333693 | 19193 | 59.9 | 0 | 0 | 0 | 2690 |
| 2018 | Woodbury | 0.440865 | 0.405133 | 19193 | 59.9 | 0 | 0 | 0 | 2690 |
| 2019 | Woodbury | 0.475018 | 0.425792 | 19193 | 59.9 | 0 | 0 | 0 | 2690 |
| 2020 | Woodbury | 0.545953 | 0.517651 | 19193 | 59.9 | 0 | 0 | 1 | 2690 |
| 2021 | Woodbury | 0.503257 | 0.482397 | 19193 | 59.9 | 0 | 0 | 1 | 2690 |
| 2022 | Woodbury | 0.500812 | 0.476711 | 19193 | 59.9 | 0 | 0 | 1 | 2690 |
| 2023 | Woodbury | 0.480536 | 0.457703 | 19193 | 59.9 | 0 | 0 | 1 | 2690 |
| 2024 | Woodbury | 0.491411 | 0.457572 | 19193 | 59.9 | 0 | 0 | 1 | 2690 |
| 2017 | Worth | 0.373333 | 0.313333 | 19195 | 53.5 | 1 | 1 | 0 | 103 |
| 2018 | Worth | 0.35503 | 0.291925 | 19195 | 53.5 | 1 | 1 | 0 | 103 |
| 2019 | Worth | 0.257862 | 0.278481 | 19195 | 53.5 | 1 | 1 | 0 | 103 |
| 2020 | Worth | 0.36076 | 0.335443 | 19195 | 53.5 | 1 | 1 | 1 | 103 |
| 2021 | Worth | 0.333333 | 0.323171 | 19195 | 53.5 | 1 | 1 | 1 | 103 |
| 2022 | Worth | 0.4 | 0.301282 | 19195 | 53.5 | 1 | 1 | 1 | 103 |
| 2023 | Worth | 0.367742 | 0.291667 | 19195 | 53.5 | 1 | 1 | 1 | 103 |
| 2024 | Worth | 0.33758 | 0.333333 | 19195 | 53.5 | 1 | 1 | 1 | 103 |
| 2017 | Wright | 0.272455 | 0.224784 | 19197 | 59.2 | 1 | 0 | 0 | 239 |
| 2018 | Wright | 0.294833 | 0.281346 | 19197 | 59.2 | 1 | 0 | 0 | 239 |
| 2019 | Wright | 0.28125 | 0.244957 | 19197 | 59.2 | 1 | 0 | 0 | 239 |
| 2020 | Wright | 0.467836 | 0.379947 | 19197 | 59.2 | 1 | 0 | 1 | 239 |
| 2021 | Wright | 0.444444 | 0.342037 | 19197 | 59.2 | 1 | 0 | 1 | 239 |
| 2022 | Wright | 0.422857 | 0.336 | 19197 | 59.2 | 1 | 0 | 1 | 239 |
| 2023 | Wright | 0.398329 | 0.31044 | 19197 | 59.2 | 1 | 0 | 1 | 239 |
| 2024 | Wright | 0.340361 | 0.314286 | 19197 | 59.2 | 1 | 0 | 1 | 239 |

**Supplemental Exhibit 3.1 – County Classifications**

|  | **Above Median COVID Vaccination Rate** | **Below Median COVID Vaccination Rate** | |
| --- | --- | --- | --- |
| **Urban** | Benton  Black Hawk  Bremer  Dallas  Dubuque  Grundy  Guthrie  Johnson  Jones  Linn  Madison  Mills  Polk  Pottawattamie  Scott  Story  Warren  Washington  Woodbury | Harrison  Plymouth | |
| **Rural** | Adams  Allamakee  Audubon  Boone  Buena Vista  Butler  Calhoun  Carroll  Cass  Cedar  Cerro Gordo  Chickasaw  Clarke  Clinton  Dickinson  Greene  Hamilton  Hardin  Iowa  Jasper  Marshall  Montgomery  Muscatine  Page  Poweshiek  Shelby  Tama  Webster  Winneshiek  Wright | Adair  Appanoose  Buchanan  Cherokee  Clay  Clayton  Crawford  Davis  Decatur  Delaware  Des Moines  Emmet  Fayette  Floyd  Franklin  Fremont  Hancock  Henry  Howard  Humboldt  Ida  Jackson  Jefferson  Keokuk | Kossuth  Lee  Louisa  Lucas  Lyon  Mahaska  Marion  Mitchell  Monona  Monroe  O’Brien  Osceola  Palo Alto  Pocahontas  Ringgold  Sac  Sioux  Taylor  Union  Van Buren  Wapello  Wayne  Winnebago  Worth |

**Supplemental Exhibit 3.2 – Pre/Post Pandemic HPV Completion Rates, by county**

|  | **Female HPV Vaccine Completion** | | **Male HPV Vaccine Completion** | |
| --- | --- | --- | --- | --- |
| **County** | **2017-2019** | **2021-2024** | **2017-2019** | **2021-2024** |
| Adair | 0.68 | 0.59 | 0.61 | 0.57 |
| Adams | 0.45 | 0.60 | 0.38 | 0.54 |
| Allamakee | 0.38 | 0.43 | 0.31 | 0.36 |
| Appanoose | 0.29 | 0.33 | 0.19 | 0.26 |
| Audubon | 0.50 | 0.57 | 0.49 | 0.46 |
| Benton | 0.57 | 0.58 | 0.48 | 0.57 |
| Black Hawk | 0.53 | 0.54 | 0.45 | 0.54 |
| Boone | 0.47 | 0.54 | 0.46 | 0.52 |
| Bremer | 0.57 | 0.60 | 0.53 | 0.59 |
| Buchanan | 0.51 | 0.52 | 0.44 | 0.51 |
| Buena Vista | 0.24 | 0.31 | 0.21 | 0.25 |
| Butler | 0.58 | 0.62 | 0.46 | 0.54 |
| Calhoun | 0.39 | 0.50 | 0.31 | 0.44 |
| Carroll | 0.35 | 0.40 | 0.27 | 0.33 |
| Cass | 0.52 | 0.55 | 0.53 | 0.52 |
| Cedar | 0.45 | 0.56 | 0.43 | 0.50 |
| Cerro Gordo | 0.44 | 0.46 | 0.42 | 0.42 |
| Cherokee | 0.39 | 0.47 | 0.35 | 0.38 |
| Chickasaw | 0.44 | 0.52 | 0.41 | 0.40 |
| Clarke | 0.45 | 0.46 | 0.35 | 0.37 |
| Clay | 0.37 | 0.39 | 0.34 | 0.30 |
| Clayton | 0.38 | 0.42 | 0.28 | 0.38 |
| Clinton | 0.33 | 0.30 | 0.27 | 0.28 |
| Crawford | 0.34 | 0.42 | 0.26 | 0.39 |
| Dallas | 0.50 | 0.55 | 0.46 | 0.54 |
| Davis | 0.17 | 0.26 | 0.14 | 0.28 |
| Decatur | 0.26 | 0.34 | 0.21 | 0.35 |
| Delaware | 0.50 | 0.53 | 0.45 | 0.46 |
| Des Moines | 0.25 | 0.27 | 0.18 | 0.23 |
| Dickinson | 0.46 | 0.49 | 0.46 | 0.47 |
| Dubuque | 0.29 | 0.41 | 0.26 | 0.37 |
| Emmet | 0.33 | 0.39 | 0.28 | 0.31 |
| Fayette | 0.45 | 0.53 | 0.38 | 0.48 |
| Floyd | 0.30 | 0.42 | 0.30 | 0.33 |
| Franklin | 0.36 | 0.40 | 0.31 | 0.38 |
| Fremont | 0.38 | 0.37 | 0.33 | 0.37 |
| Greene | 0.49 | 0.55 | 0.42 | 0.48 |
| Grundy | 0.49 | 0.59 | 0.43 | 0.59 |
| Guthrie | 0.54 | 0.55 | 0.53 | 0.52 |
| Hamilton | 0.42 | 0.45 | 0.31 | 0.36 |
| Hancock | 0.37 | 0.40 | 0.33 | 0.38 |
| Hardin | 0.39 | 0.42 | 0.37 | 0.37 |
| Harrison | 0.44 | 0.40 | 0.38 | 0.45 |
| Henry | 0.28 | 0.32 | 0.24 | 0.27 |
| Howard | 0.46 | 0.45 | 0.42 | 0.39 |
| Humboldt | 0.52 | 0.54 | 0.45 | 0.47 |
| Ida | 0.34 | 0.32 | 0.28 | 0.31 |
| Iowa | 0.39 | 0.48 | 0.34 | 0.43 |
| Jackson | 0.23 | 0.33 | 0.20 | 0.23 |
| Jasper | 0.38 | 0.44 | 0.30 | 0.45 |
| Jefferson | 0.30 | 0.35 | 0.21 | 0.29 |
| Johnson | 0.43 | 0.49 | 0.40 | 0.47 |
| Jones | 0.44 | 0.56 | 0.37 | 0.52 |
| Keokuk | 0.43 | 0.49 | 0.40 | 0.45 |
| Kossuth | 0.48 | 0.49 | 0.41 | 0.46 |
| Lee | 0.36 | 0.44 | 0.31 | 0.39 |
| Linn | 0.46 | 0.55 | 0.42 | 0.51 |
| Louisa | 0.35 | 0.44 | 0.34 | 0.36 |
| Lucas | 0.38 | 0.40 | 0.34 | 0.35 |
| Lyon | 0.31 | 0.39 | 0.24 | 0.34 |
| Madison | 0.55 | 0.55 | 0.47 | 0.56 |
| Mahaska | 0.34 | 0.37 | 0.26 | 0.33 |
| Marion | 0.37 | 0.41 | 0.30 | 0.37 |
| Marshall | 0.55 | 0.58 | 0.52 | 0.53 |
| Mills | 0.34 | 0.35 | 0.30 | 0.32 |
| Mitchell | 0.42 | 0.45 | 0.40 | 0.43 |
| Monona | 0.57 | 0.52 | 0.51 | 0.47 |
| Monroe | 0.38 | 0.38 | 0.32 | 0.29 |
| Montgomery | 0.39 | 0.43 | 0.29 | 0.41 |
| Muscatine | 0.41 | 0.45 | 0.35 | 0.41 |
| O’Brien | 0.47 | 0.51 | 0.41 | 0.47 |
| Osceola | 0.35 | 0.48 | 0.37 | 0.46 |
| Page | 0.43 | 0.50 | 0.43 | 0.47 |
| Palo Alto | 0.38 | 0.39 | 0.33 | 0.33 |
| Plymouth | 0.42 | 0.46 | 0.34 | 0.44 |
| Pocahontas | 0.47 | 0.50 | 0.38 | 0.49 |
| Polk | 0.47 | 0.53 | 0.42 | 0.51 |
| Pottawattamie | 0.41 | 0.44 | 0.37 | 0.43 |
| Poweshiek | 0.51 | 0.47 | 0.50 | 0.46 |
| Ringgold | 0.51 | 0.55 | 0.43 | 0.51 |
| Sac | 0.37 | 0.45 | 0.30 | 0.35 |
| Scott | 0.38 | 0.39 | 0.35 | 0.36 |
| Shelby | 0.42 | 0.47 | 0.46 | 0.45 |
| Sioux | 0.33 | 0.38 | 0.31 | 0.34 |
| Story | 0.46 | 0.47 | 0.43 | 0.46 |
| Tama | 0.50 | 0.57 | 0.49 | 0.55 |
| Taylor | 0.46 | 0.44 | 0.43 | 0.47 |
| Union | 0.51 | 0.56 | 0.43 | 0.51 |
| Van Buren | 0.26 | 0.25 | 0.24 | 0.20 |
| Wapello | 0.39 | 0.43 | 0.35 | 0.39 |
| Warren | 0.46 | 0.56 | 0.41 | 0.51 |
| Washington | 0.37 | 0.43 | 0.33 | 0.41 |
| Wayne | 0.38 | 0.38 | 0.34 | 0.31 |
| Webster | 0.41 | 0.49 | 0.34 | 0.44 |
| Winnebago | 0.31 | 0.36 | 0.29 | 0.31 |
| Winneshiek | 0.54 | 0.60 | 0.48 | 0.57 |
| Woodbury | 0.43 | 0.49 | 0.39 | 0.47 |
| Worth | 0.33 | 0.36 | 0.29 | 0.31 |
| Wright | 0.28 | 0.40 | 0.25 | 0.33 |

**Supplemental Exhibit 3.3 –Post-COVID-19 Pandemic Year-by-Year Changes in HPV Vaccine Completion Rates**

| **Year** | **Group** | **Sex** | **Estimate** | **SE** | **Lower** | **Upper** |
| --- | --- | --- | --- | --- | --- | --- |
| 2021 | Rural-Low | Female | 0.028 | 0.005 | 0.018 | 0.038 |
| 2022 | Rural-Low | Female | 0.022 | 0.007 | 0.008 | 0.036 |
| 2023 | Rural-Low | Female | 0.022 | 0.007 | 0.008 | 0.036 |
| 2024 | Rural-Low | Female | 0.015 | 0.008 | -0.001 | 0.031 |
| 2021 | Rural-Low | Male | 0.036 | 0.006 | 0.024 | 0.048 |
| 2022 | Rural-Low | Male | 0.032 | 0.007 | 0.018 | 0.046 |
| 2023 | Rural-Low | Male | 0.022 | 0.007 | 0.008 | 0.036 |
| 2024 | Rural-Low | Male | 0.015 | 0.007 | 0.001 | 0.029 |
| 2021 | Rural-High | Female | 0.037 | 0.006 | 0.025 | 0.049 |
| 2022 | Rural-High | Female | 0.037 | 0.006 | 0.025 | 0.049 |
| 2023 | Rural-High | Female | 0.034 | 0.009 | 0.016 | 0.052 |
| 2024 | Rural-High | Female | 0.041 | 0.013 | 0.016 | 0.066 |
| 2021 | Rural-High | Male | 0.041 | 0.007 | 0.027 | 0.055 |
| 2022 | Rural-High | Male | 0.026 | 0.008 | 0.010 | 0.042 |
| 2023 | Rural-High | Male | 0.019 | 0.010 | -0.001 | 0.039 |
| 2024 | Rural-High | Male | 0.026 | 0.013 | 0.001 | 0.051 |
| 2021 | Urban-High | Female | 0.037 | 0.005 | 0.027 | 0.047 |
| 2022 | Urban-High | Female | 0.036 | 0.006 | 0.024 | 0.048 |
| 2023 | Urban-High | Female | 0.031 | 0.008 | 0.015 | 0.047 |
| 2024 | Urban-High | Female | 0.030 | 0.008 | 0.014 | 0.046 |
| 2021 | Urban-High | Male | 0.059 | 0.006 | 0.047 | 0.071 |
| 2022 | Urban-High | Male | 0.051 | 0.006 | 0.039 | 0.063 |
| 2023 | Urban-High | Male | 0.048 | 0.006 | 0.036 | 0.060 |
| 2024 | Urban-High | Male | 0.047 | 0.006 | 0.035 | 0.059 |
| 2021 | Urban-Low | Female | 0.014 | 0.028 | -0.041 | 0.069 |
| 2022 | Urban-Low | Female | -0.001 | 0.033 | -0.066 | 0.064 |
| 2023 | Urban-Low | Female | -0.022 | 0.021 | -0.063 | 0.019 |
| 2024 | Urban-Low | Female | -0.029 | 0.032 | -0.092 | 0.034 |
| 2021 | Urban-Low | Male | 0.091 | 0.032 | 0.028 | 0.154 |
| 2022 | Urban-Low | Male | 0.070 | 0.017 | 0.037 | 0.103 |
| 2023 | Urban-Low | Male | 0.031 | 0.007 | 0.017 | 0.045 |
| 2024 | Urban-Low | Male | 0.018 | 0.020 | -0.021 | 0.057 |

**Supplemental Exhibit 3.4 –Post-COVID-19 Pandemic Year-by-Year Changes in HPV Vaccine Completion Rates**

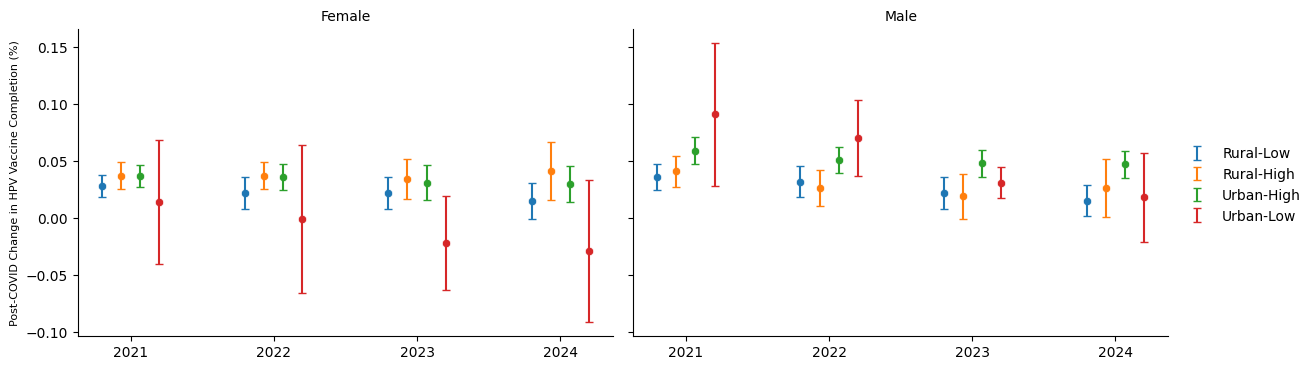

**Supplemental Exhibit 3.5 – Trends in HPV Vaccine Completion Rates – excluding year 2019-2020**

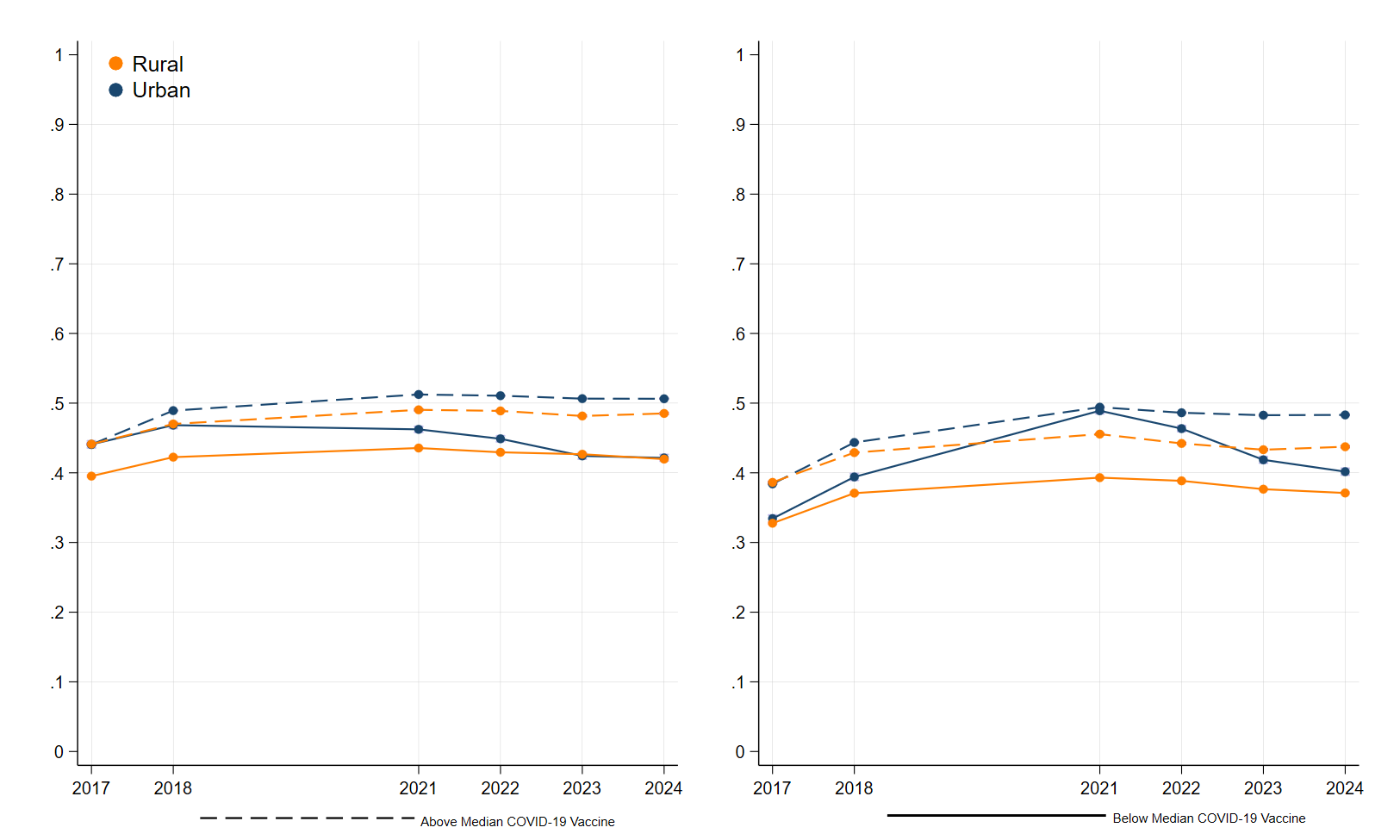

**Supplemental Exhibit 6 – Estimated Association Between COVID-19 Pandemic and HPV Vaccination Completion Rates (Females) – excluding years 2019-2020**

|  | (1) | (2) | (3) | (4) |
| --- | --- | --- | --- | --- |
| Post | 0.038***  [0.026, 0.050] | 0.043***  [0.025, 0.060] | 0.043***  [0.027, 0.056] | 0.044***  [0.026, 0.061] |
| Post # Rural |  | -0.014  [-0.034, 0.006] |  | -0.010  [-0.034, 0.015] |
| Post # Below COVID |  |  | -0.020*  [-0.039, -0.002] | -0.057  [-0.128, -.014] |
| Post # Rural # Below COVID |  |  |  | 0.046  [-0.028, 0.121] |

* p<0.05, ** p < 0.01, *** p < 0.001

**Supplemental Exhibit 7 – Estimated Association Between COVID-19 Pandemic and HPV Vaccination Completion Rates (Males) - excluding years 2019-2020**

|  | (1) | (2) | (3) | (4) |
| --- | --- | --- | --- | --- |
| Post | 0.060***  [0.048, 0.073] | 0.073***  [0.058, 0.087] | 0.065***  [-0.024, -0.005] | 0.073***  [0.058, 0.087] |
| Post # Rural |  | -0.034***  [-0.053, -0.016] |  | -0.034**  [-.0.060, -0.009] |
| Post # Below COVID |  |  |  | 0.011  [-0.039, 0.062] |
| Post # Rural # Below COVID |  |  |  | -0.011  [-0.067, 0.045] |

* p<0.05, ** p < 0.01, *** p < 0.00
